# Statin uses and skeletal muscle-related phenotypes: insights from epidemiological and Mendelian randomization analyses

**DOI:** 10.1101/2024.09.16.24313777

**Authors:** Fan Tang, Zhanchao Chen, Hongbing Qiu, Yige Liu, Yanjiao Shen, Yiying Zhang, Shanjie Wang, Bo Yu

## Abstract

**Background:** The association between statin use and skeletal muscle-related side effects is always controversial. This study aimed to comprehensively investigate the associations between statin use and muscle-related phenotypes including sarcopenia, sarcopenic obesity, serum lactate dehydrogenase (LDH), and musculoskeletal pain symptoms among adults with indications for statin use for secondary prevention (cardiovascular disease, diabetes, or hyperlipidemia).

**Methods:** This cross-sectional study included 22,549 patients aged ≥20 years with cardiovascular disease, diabetes, or hyperlipidemia. Weighted generalized linear regression analysis and propensity score matching methods were used to estimate the associations between the use of statins or other lipid-lowering agents and skeletal muscle-related phenotypes. Mendelian randomization (MR) analysis was additionally used to verify the causal relationship between statin use and skeletal muscle-related phenotypes.

**Results:** The weighted mean age was 59 years, 50.3% were male, and 37.6% (n=8,481) received statin treatment. In the unadjusted model, compared with adults without any lipid-lowering drugs, statin use was associated with a higher likelihood of sarcopenia (appendicular skeletal muscle mass [ASM]/Body mass index [BMI] OR 1.35 (95%CI 1.12 to 1.62, p < 0.001), ASM/weight [Wt] OR 1.86 (95%CI 1.62 to 2.13, p < 0.001), max HGS β -3.01 (95% CI -3.97 to -2.06, p < 0.001), relative HGS β -0.23 (95% CI -0.30 to -0.17, p < 0.001) and combined HGS β -5.90 (95% CI -7.86 to -3.93, p < 0.001)), sarcopenic obesity (ASM/height squared [Ht^2^] and body fat percentage definition [OR 1.36 (95% CI 1.13 to 1.63, p < 0.001]). After multivariable adjustment or propensity score match, the independent associations of statin use with sarcopenia, sarcopenic obesity, HGS, LDH, and musculoskeletal pain became nonsignificant. Stepwise regression suggested that age was the predominant confounding factor for the associations. MR analysis also revealed no significant causality between statin use and skeletal muscle-related phenotypes.

**Conclusions:** Our epidemiological and MR analyses did not support the causality between statin use and skeletal muscle-related phenotypes. A higher likelihood of skeletal muscle-related adverse phenotypes in statin users may be attributed to age. Future studies should further explore the biological factors that may affect statin-related muscle phenotypes to provide evidence for the safety of statins.

## 1. Introduction

Statins are the first-line prescribed drugs that lower lipid levels by inhibiting 3-hydroxy-3-methylglutaryl-CoA reductase[1]. Numerous studies demonstrated the cornerstone status of statins in preventing the risk of atherosclerotic cardiovascular disease (ASCVD) and relevant mortality[2, 3]. Thus, current clinical guidelines underline the importance of statin use in cardiovascular disease, diabetes, and hyperlipidemia[2, 4, 5].

Despite the significant benefits of statins, the controversy surrounding Statin-associated muscle symptoms (SAMS) persists. The potential adverse effects of statin use remain a significant subject of discussion, prompting concerns among patients and physicians [6, 7]. SAMS, encompassing muscle-related symptoms such as myalgia, tenderness, stiffness, cramps, and weakness, with or without elevated creatine kinase levels, are frequently reported as the predominant side effects. These symptoms can lead to causes medication nonadherence, potentially leading to suboptimal cardiovascular management [8, 9]. Statin discontinuation related to SAMS has been shown to cause a nearly 3-fold increased risk of cardiovascular events[10]. Most randomized controlled trials indicate that the risk of SAMS is minimal or insignificant compared with a placebo[11]. A study showed[2]that older adults receiving statins for primary and secondary prevention do not have an increased risk of myopathy. A recent systematic review showed[12]that statin use was not significantly related to the risk of SAMS and relevant mortality among older statin users. A cross-sectional study showed no correlation between statin use and musculoskeletal pain in arthritis patients[13]. However, it is important to note that previous trials and meta-analyses may have several limitations. For example, study points by self-reported symptoms may be subjective; severe myopathy and rhabdomyolysis, both are rare in humans as statin-related musculoskeletal disorders; and the study population may lack representativeness due to small sample sizes and potential selection bias[12].

Sarcopenia is an age-related syndrome characterized by loss of muscle mass, strength, and function[14]. Dual-energy X-ray absorptiometry (DXA) is employed for assessing total body lean tissue mass or appendicular skeletal muscle mass[15]. However, the relationship between statins and muscle mass or sarcopenia has not been reported. Handgrip strength (HGS) serves as another indicator for assessing skeletal muscle function[16]. Only two studies reported inconsistent results on the association between statins and muscle function (e.g., grip strength, and gait speed) in older adults[17, 18]. Overall, there’s a lack of large-scale studies comprehensively examining the association between statin use and instrument-based muscle characteristics. Mendelian randomization (MR) analysis is acknowledged as a complementary method to imitate randomized controlled trials, which is increasingly recognized as an emerging epidemiological approach that utilizes genetic factors to evaluate the causal impact of exposure on a particular outcome[19]. Therefore, this study combined a large-scale observational study in the National Nutrition Health and Nutrition Examination Survey (1999-2020) 1999–2020, and a two-sample MR analysis was conducted to evaluate the relationships between statin use and muscle-related phenotypes including sarcopenia, sarcopenic obesity, serum lactate dehydrogenase (LDH), and musculoskeletal pain among adults with the indications for statin use for secondary prevention (cardiovascular disease, diabetes, or hyperlipidemia).

## 2. Methods

### 2.1 Study Population

As we have previously reported[20], the National Nutrition Health and Nutrition Examination Survey employs a stratified, multistage probability sampling method to represent the noninstitutionalized population of the United States. In this study, datasets from 11 two-year cycles of the National Nutrition Health and Nutrition Examination Survey (from 1999–2000 to 2017–2020). The data from the 11 cycles were standardized and combined, using interview weights consistent with the recommendations from the National Center for Health Statistics.

A total of 116,876 participants were involved in the survey over two cycles. We excluded individuals aged < 20 years (n = 52,563), those without assessment for cardiovascular disease (CVD), diabetes, or hyperlipidemia (n = 37,051), and those lacking data on muscle-related phenotypes (n = 4,713). In total, 22,549 subjects were analyzed, including 13,474 non-statin users, 8,481 statin users, and 594 users of lipid-lowering drugs other than statins (**Figure 1**). The study was approved by the research ethics review board of the Centers for Disease Control and Prevention, and all participants provided informed consent.

**Figure 1.**
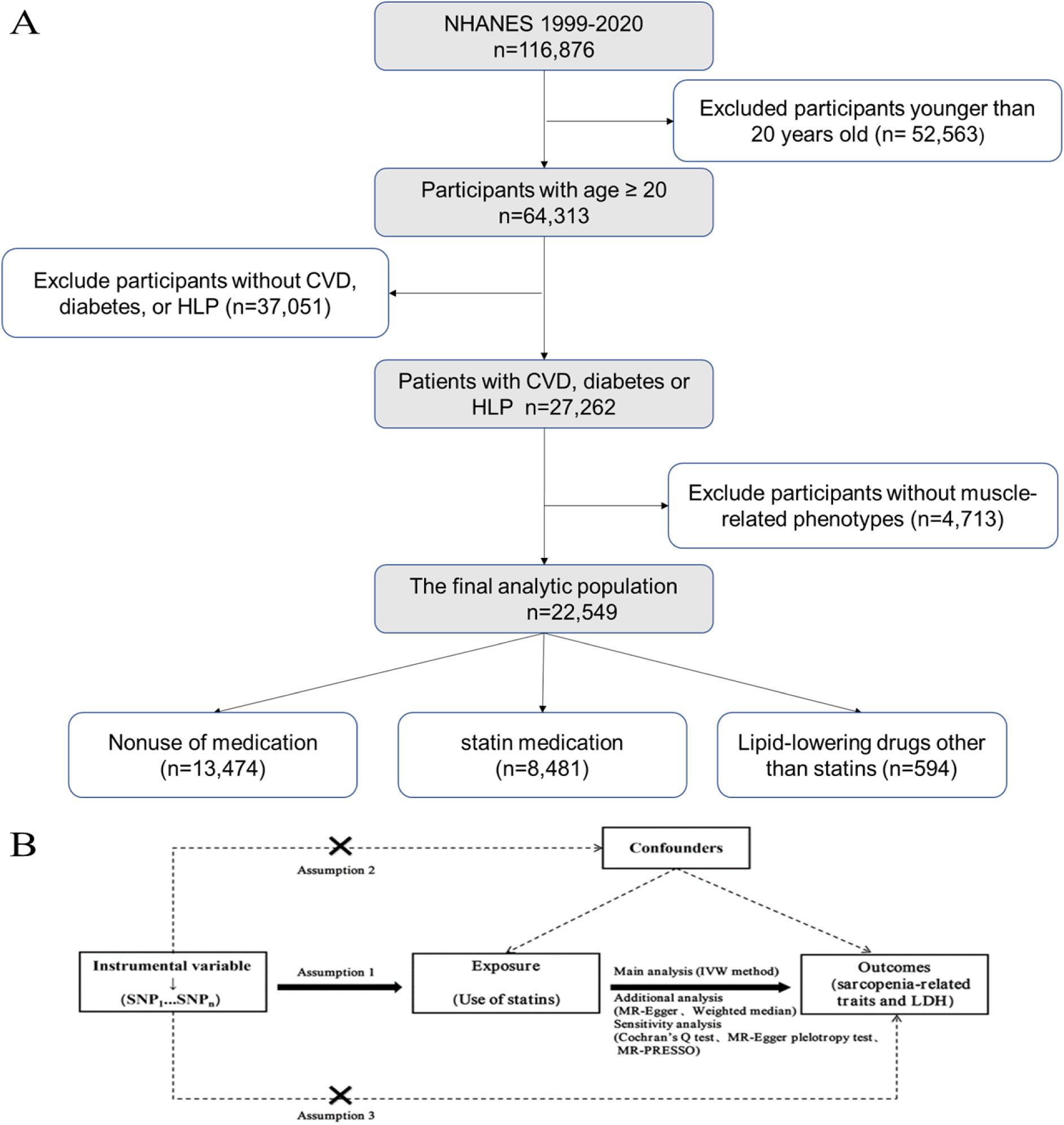
Flow of study. Abbreviations: NHANES, National Health and Nutrition Examination Survey; CVD, cardiovascular disease; HLP, hyperlipidemia; SNP, Single Nucleotide Polymorphism; LDH, Lactate Dehydrogenase. A: Flow of study. B: Two-sample Mendelian randomization study design. Arrows represent associations. Dashed lines with “×” in the middle represent genetic variants applied as instrumental variables for Statins that should not be associated with potential confounding factors and do not exert an effect on sarcopenia-related traits and LDH via other pathways in assumptions 2 and 3, respectively. Abbreviations: SNP, Single nucleotide polymorphism; IVW, Inverse variance weighted; MR-PRESSO, MR-pleiotropy residual sum and outlier; LDH, Lactic dehydrogenase.

CVD was defined as a self-reported history of coronary heart disease, heart failure, or stroke. Diabetes was defined as a physician-reported diagnosis, plasma HbA1c ≥ 6.5%, or fasting blood glucose ≥ 7.0 mmol/L[21]. Hyperlipidemia was defined as a physician-reported diagnosis in the primary analysis. In sensitivity analyses, hyperlipidemia is defined as total cholesterol 200 mg/dL, triglycerides 150 mg/dL, HDL 40 mg/dL in men, 50 mg/dL in women, or hypolipidemia—high-density lipoprotein cholesterol 130 mg/dL[22]. Additionally, individuals reporting the use of cholesterol-lowering medications were also categorized as having hyperlipidemia.

Two-sample MR was performed to evaluate the causal effects between s statin use and muscle-related phenotypes (**Figure 1**). The current MR research is based on three main assumptions[23]. First, the selected SNPs were associated with statins; second, the SNPs were not associated with other confounding factors; third, the SNPs only affected sarcopenia-related indicators and the risk of LDH through statins, without affecting other pathways. Detailed information about the data downloading and screening processes is displayed in **Table S1** and **Table S2.**

### 2.2 The use of statins and lipid-lowering agents

At baseline, during a personal interview, participants were asked about the prescription medications they had taken in the past 30 days. The prescription drug names are recorded on the drug container label. Each drug is associated with a standardized generic prescription drug name[20]. **Table S3** contains a list of statins and lipid-lowering drugs.

### 2.3 Definition of skeletal muscle-related phenotypes

#### 2.3.1 Sarcopenia and sarcopenic obesity

The survey provides an accurate set of muscle mass data, assessed by dual-energy X-ray absorptiometry (DEXA). Appendicular skeletal muscle mass (ASM (kg)) was defined as the sum of the muscle mass of the four limbs. The skeletal muscle index was calculated as the appendicular skeletal muscle mass divided by the body mass index (BMI) or weight (Wt). In this study, the cutoff value for sarcopenia was defined by BMI-adjusted ASM according to the Foundation for National Institutes of Health (FNIH) criteria[24], male < 0.789, female < 0.512. The cutoff for sarcopenia was defined by a weight-adjusted ASM of 29.76% for males and 22.31% for females[25]. The appendicular skeletal mass index (ASMI) was defined as the ASM divided by the square of the height(ht^2^). According to the European Working Group on Sarcopenia in Older Population(EWGSOP2)[26], sarcopenia was defined as an ASMI <7 kg/m^2^ in men or <5.5 kg/m^2^ in women[27]. Subjects were classified as obese using standard BMI categories (≥30 kg/m^2^) or a body fat percentage ≥25% for men and ≥35% for women. If the subject met the criteria for both sarcopenia and obesity using these definitions, a diagnosis of sarcopenic obesity was considered.

The muscle strength/grip strength was assessed by isometric HGS using a handgrip dynamometer[28]. Grip strength testing is typically conducted with participants in a standing position unless they have physical constraints. Muscle strength was measured in three indicators: maximum HGS, combined HGS, and relative HGS. Participants underwent three HGS tests for both their left and right hands. The study selected the highest value from the three tests for maximum HGS; combined HGS was calculated as the sum of the highest readings from each hand. Relative grip strength was defined as the sum of the highest readings from each hand, divided by BMI.

#### 2.3.2 Lactate dehydrogenase

The survey measures serum LDH levels at baseline in all adult participants using an enzymatic rate method; more information on LDH analysis techniques can be found in the Laboratory Procedure Manual[29]

#### 2.3.3 Musculoskeletal pain in different anatomical regions

Participants were asked, ‘In the past month, have you had problems with pain that lasted for more than 24 hours?’ Those who answered ‘yes’ were asked to specify the location of the pain. We assessed reports of pain in the neck or upper back, upper extremities, lower back, and lower extremities. Musculoskeletal pain was defined as pain in one of these areas.

### 2.4 Other variables

The covariates included age, sex, race/ethnicity, education level, income-to-poverty ratio, marital status, alcohol use status, smoking status, physical activity, BMI, hypertension, cardiovascular disease, diabetes, hyperlipidemia, chronic kidney disease, chronic obstructive pulmonary disease, and cancer. The poverty income ratio was categorized into three categories: 1.5, 1.5-3.5, and >3.5[30]. BMI categories of <25 kg/m^2^, 25 to 30 kg/m^2^, and >30 kg/m2 were established. Hypertension, cardiovascular disease, diabetes, hyperlipidemia, chronic obstructive pulmonary disease, and cancer were defined based on self-reported prior diagnoses.

### 2.5 Propensity score matching

Propensity score matching was used to balance baseline differences in cardiovascular disease, diabetes, and hyperlipidemia between the statin and non-statin groups. Propensity scores were calculated for individuals using logistic regression, including all covariates. Paired patients were selected using the nearest neighbor matching algorithm without replacement, with a caliper size of 0.2 standard deviations of the propensity score logit[20].In addition, standardized differences were used after matching to assess the balance of covariates between the two groups; covariates with standardized differences less than 0.01 were considered well-balanced.

### 2.6 Statistical analysis

Statistical analyses were performed according to the analysis guidelines for the dataset. Because the amount of missing data in the covariates was small (<5%), we used multivariate interpolation with chained equations to handle the missing data[31]. The baseline characteristics of the participants were summarized via descriptive analysis according to their statin use status. Chi-square tests and t-tests were used to compare categorical and continuous data[30]. Weighted generalized linear regression analyses were used to estimate the associations between statin or lipid-lowering drug use and sarcopenic, sarcopenic obesity, HGS, LDH, and musculoskeletal pain. Multivariate models were adjusted for covariates included in propensity score matching. We further redefined the population with hyperlipidemia to examine the association between statin or lipid-lowering drug use and low muscle mass, grip strength, or musculoskeletal pain. Other analyses were limited to patients aged 65 years or older with cardiovascular disease, diabetes, or hyperlipidemia.

We performed two-sample MR analyses to assess the causal relationships between statin use and appendicular lean mass, hand grip strength (left and right), low hand grip strength, walking pace, and LDH. Handling missing values using multiple imputations. In the main analyses, we used a random-effects inverse variance weighted (IVW) approach to estimate causal effects. In addition, sensitivity analyses using weighted median, MR‒Egger, and MR-pleiotropic residuals and outliers (MR-PRESSO) methods were performed to assess the robustness of the IVW results. We considered two-sided p-values less than 0.05 as statistically significant. Stata (version 15.0) and R (version 4.1.2) were used for all analyses.

## 3. Results

### 3.1 Participant characteristics

This cross-sectional study included 22,549 individuals diagnosed with cardiovascular disease, diabetes, or hyperlipidemia at baseline between 1999 -2020. Overall, 37.6% of patients (n=8481) were taking statins at baseline. The mean age of all subjects was 59 ± 15 years, and 50.3% were male. Those taking statins were more likely to be older, have a greater body mass index, and were less likely to smoke (P< 0.05) (Table 1). Adults without any lipid-lowering drugs had a higher proportion of chronic diseases such as hypertension, chronic kidney disease, or cancer (P< 0.05) (**Table 1)**. We also matched statin and non-statin users separately (**Table S4**).

**Table 1.**
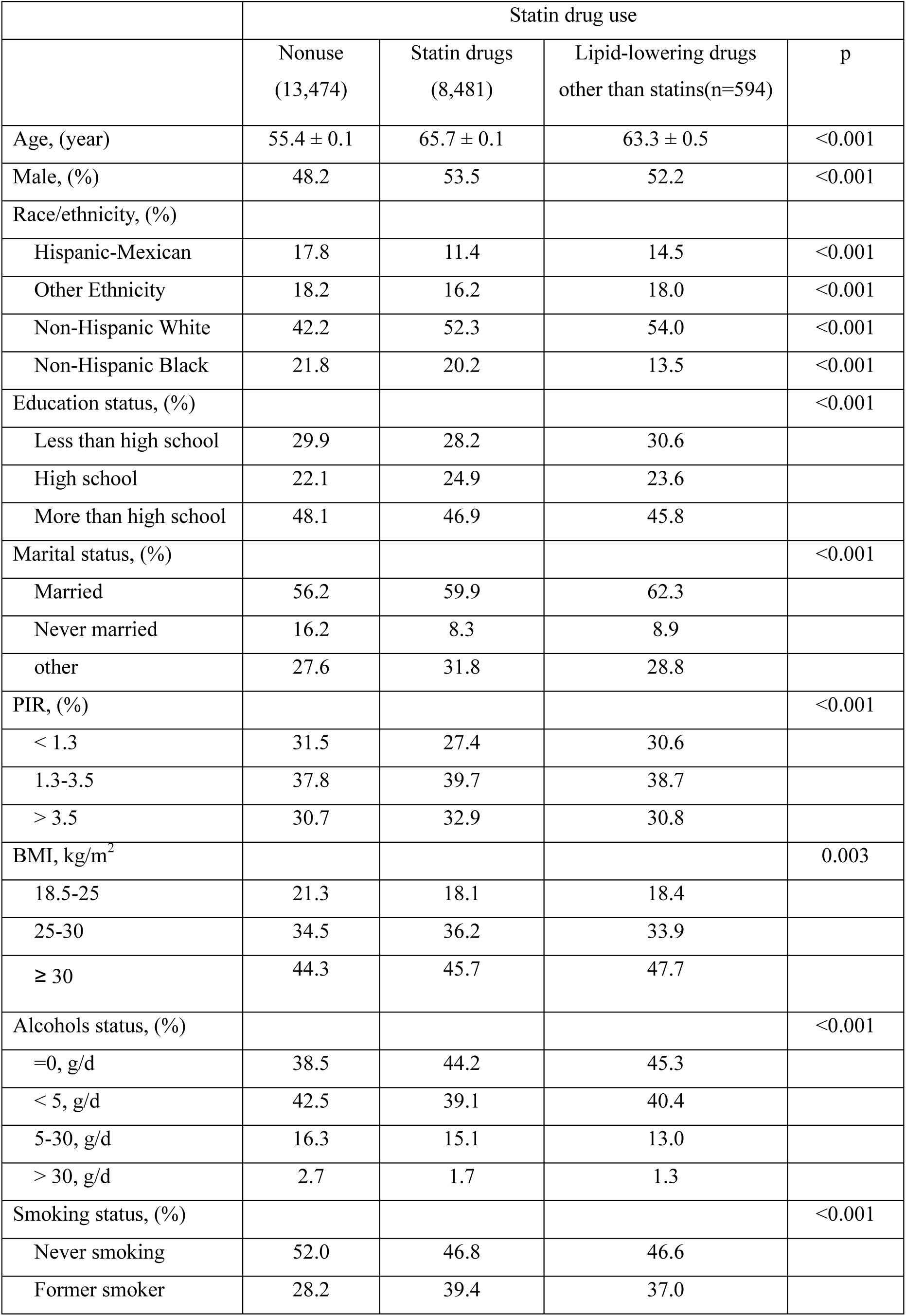
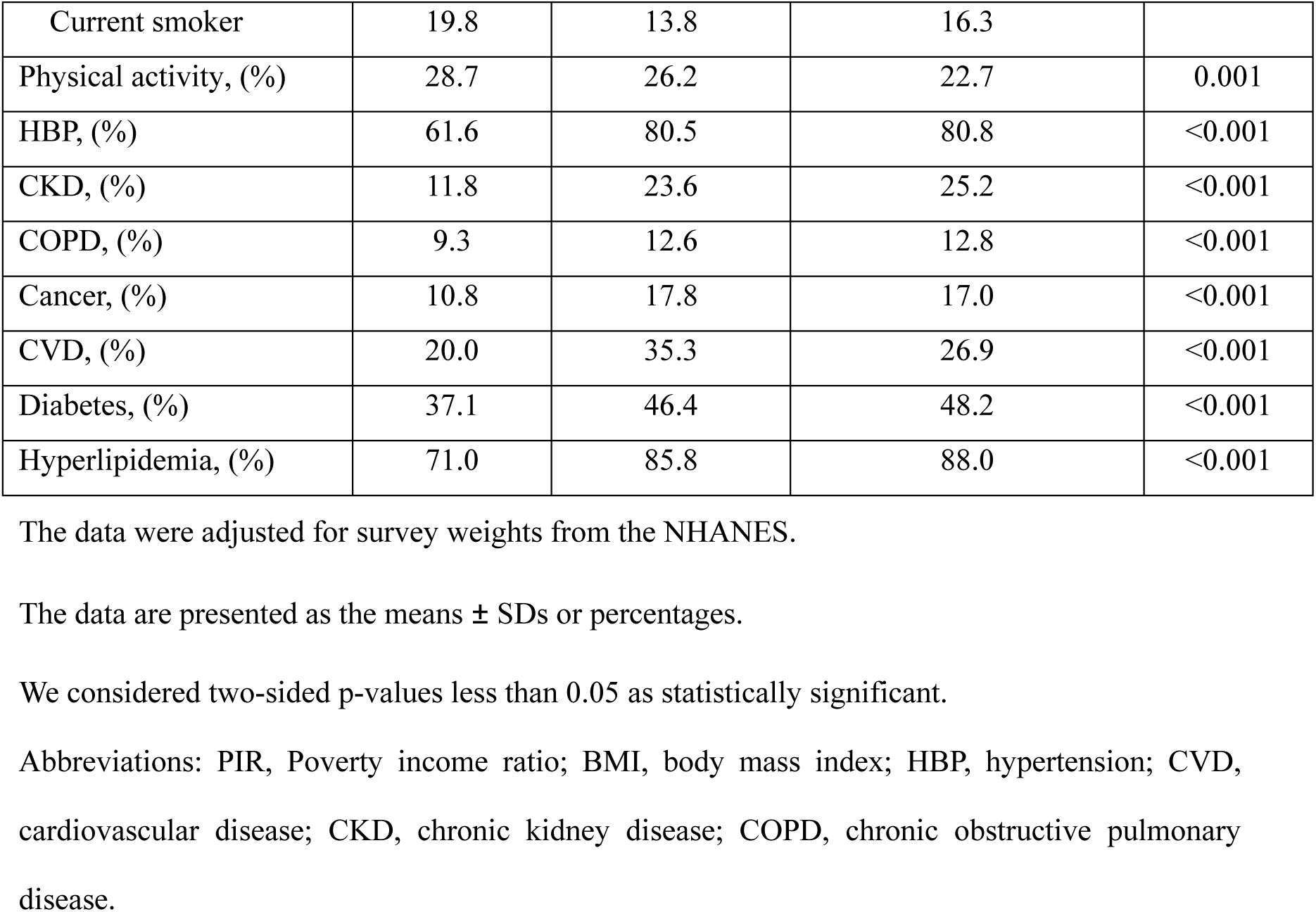
Baseline characteristics of patients with cardiovascular disease, diabetes.

### 3.2 Relationship between statin use and muscle-related phenotypes

**Table 2** shows the relationships between statin use and muscle-related phenotypes including sarcopenia, sarcopenic obesity, and LDH. Compared with adults without any lipid-lowering drugs, statin user was associated with a higher likelihood of muscle-related phenotypes, ASM/BMI (OR=1.35, 95% CI: 1.12 to 1.62), ASM/Wt (OR=1.86, 95% CI: 1.62 to 2.13), maximum HGS (β=-3.01, 95% CI: -3.97 to -2.06), relative HGS (β=-0.23 95% CI: -0.30 to -0.17), and combined HGS (β=-5.90 95% CI: -7.86 to -3.93), ASM/Ht^2^ and body fat percentage defined as sarcopenic obesity (OR=1.36, 95% CI: 1.13 to 1.63) in the unadjusted model.

**Table 2.**
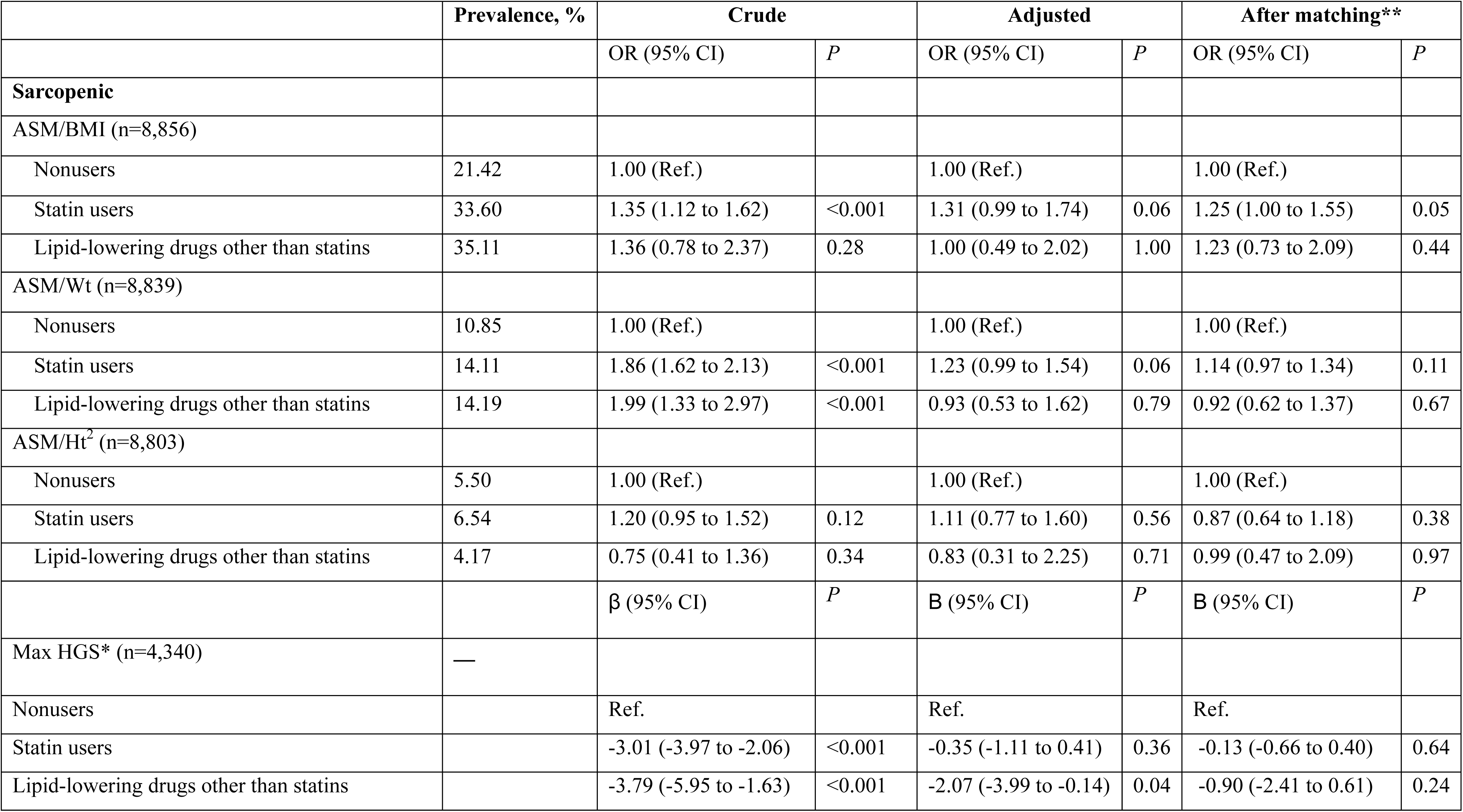
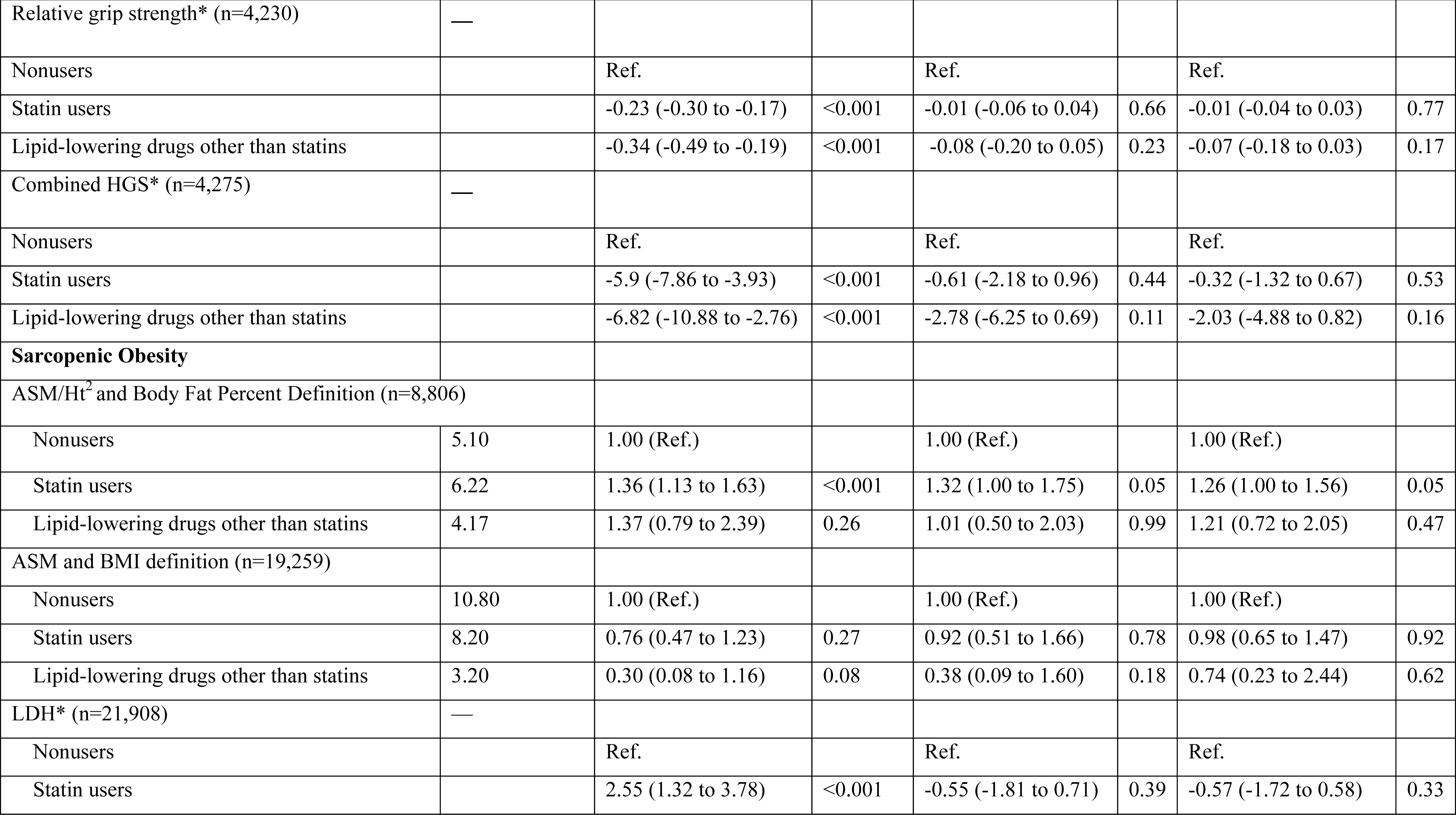
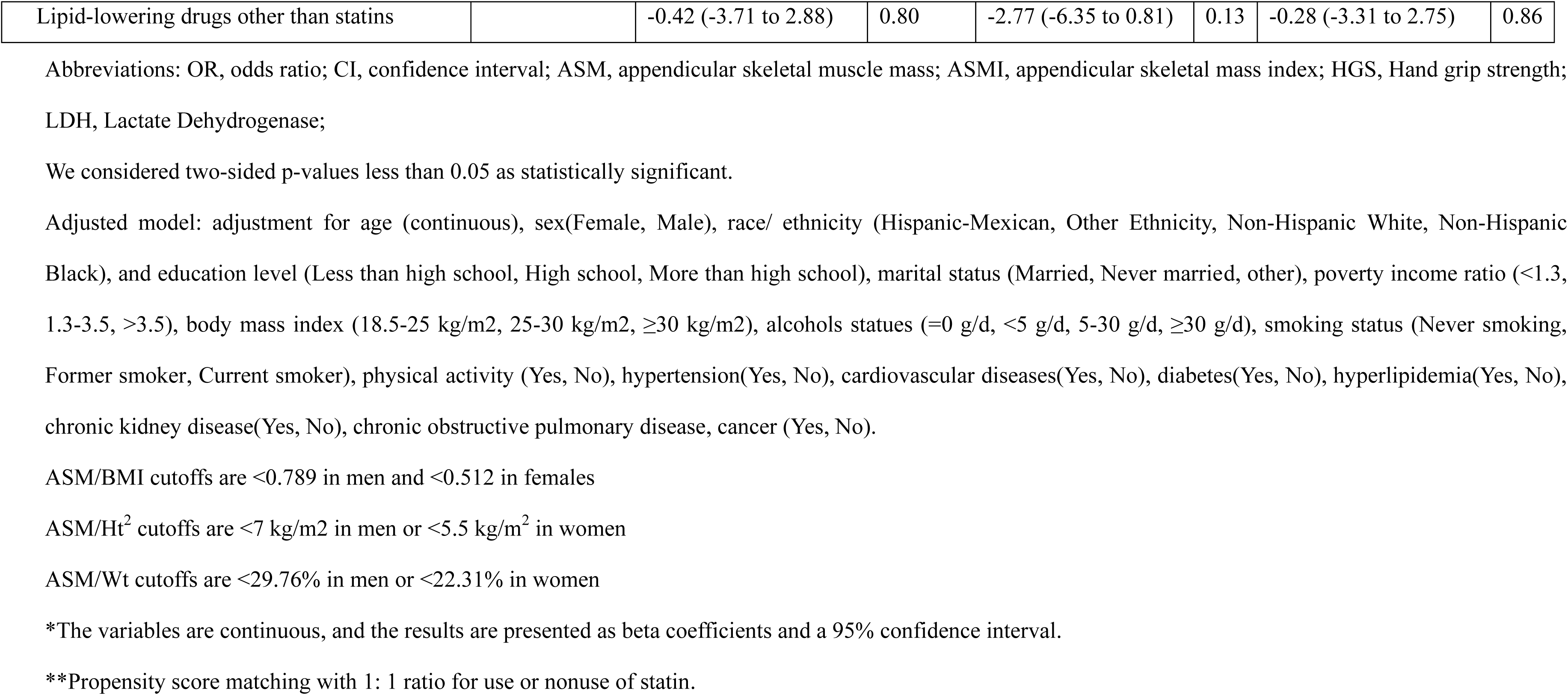
The association between statin users or lipid-lowering users and skeletal muscle-related phenotypes compared with nonusers.

Multivariate linear regression analysis revealed that compared with adults without any lipid-lowering drugs, statin user was not associated with a higher likelihood of muscle-related phenotypes including ASM/BMI (OR=1.31, 95% CI: 0.99 to 1.74), ASM/Wt (OR=1.23, 95% CI: 0.99 to 1.54), ASM/Ht^2^ (OR=1.11, 95% CI: 0.77 to 1.60), maximum HGS (β=-3.01, 95% CI: -3.97 to -2.06), relative HGS (β =-0.23 95% CI: -0.30 to -0.17), and combined HGS (β=-5.90 95% CI: -7.86 to -3.93), ASM/Ht^2^ and body fat percentage defined as sarcopenic obesity (OR= 1.32, 95% CI: 1.00 to 1.75), ASM and BMI defined as sarcopenic obesity (OR=0.92, 95% CI: 0.51 to 1.66).

We further performed propensity score-matched analyses (**Table 2**) to account for possible confounding factors that may have contributed to the protective associations between statin use and muscle-related phenotypes. Statin user was not associated with a higher likelihood of muscle-related phenotypes including ASM/BMI (OR=1.25, 95% CI: 1.00 to 1.55), ASM/Wt (OR=1.14, 95% CI: 0.97 to 1.34), ASM/Ht^2^ (OR=0.87, 95% CI: 0.64 to 1.18), maximum HGS (β=-0.13 95% CI: -0.66 to 0.40), relative HGS (β= -0.01 95% CI: -0.04 to 0.03), combined HGS (β=-0.32 95% CI: -1.32 to 0.67), or LDH (β=-0.57, 95% CI: -1.72 to 0.58), ASM/Ht^2^ and body fat percentage defined as sarcopenic obesity (OR= 1.26, 95% CI: 1.00 to 1.56), ASM and BMI defined as sarcopenic obesity (OR=0.74, 95% CI: 0.23 to 2.44).

**Table 3** shows the adjusted and unadjusted odds ratios for musculoskeletal pain and statin use. Multivariate linear regression analysis revealed that compared with without any lipid-lowering drugs, statin user was not associated with a higher likelihood of musculoskeletal pain including any regional pain (OR=1.15, 95% CI: 0.88 to 1.50), neck/upper back pain (OR= 0.96, 95% CI: 0.64 to 1.44), upper extremities pain (OR=1.11, 95% CI: 0.81 to 1.51), lower back pain (OR=1.05, 95% CI: 0.77 to 1.45) or lower extremities pain (OR=1.12, 95% CI: 0.83 to 1.51). We further performed propensity score-matched analyses (**Table 3**) to account for possible confounding factors that may have contributed to the protective association between statin use and musculoskeletal pain. Compared without any lipid-lowering drugs, statin user was not associated with any regional pain (OR=1.09, 95% CI: 0.91 to 1.31), neck/upper back pain (OR=0.94, 95% CI: 0.67 to 1.32), upper extremities pain (OR=1.13, 95% CI: 0.93 to 1.39), lower back pain (OR=1.05, 95% CI: 0.79 to 1.39) or lower extremities pain (OR=1.08, 95% CI: 0.84 to 1.41).

**Table 3.**
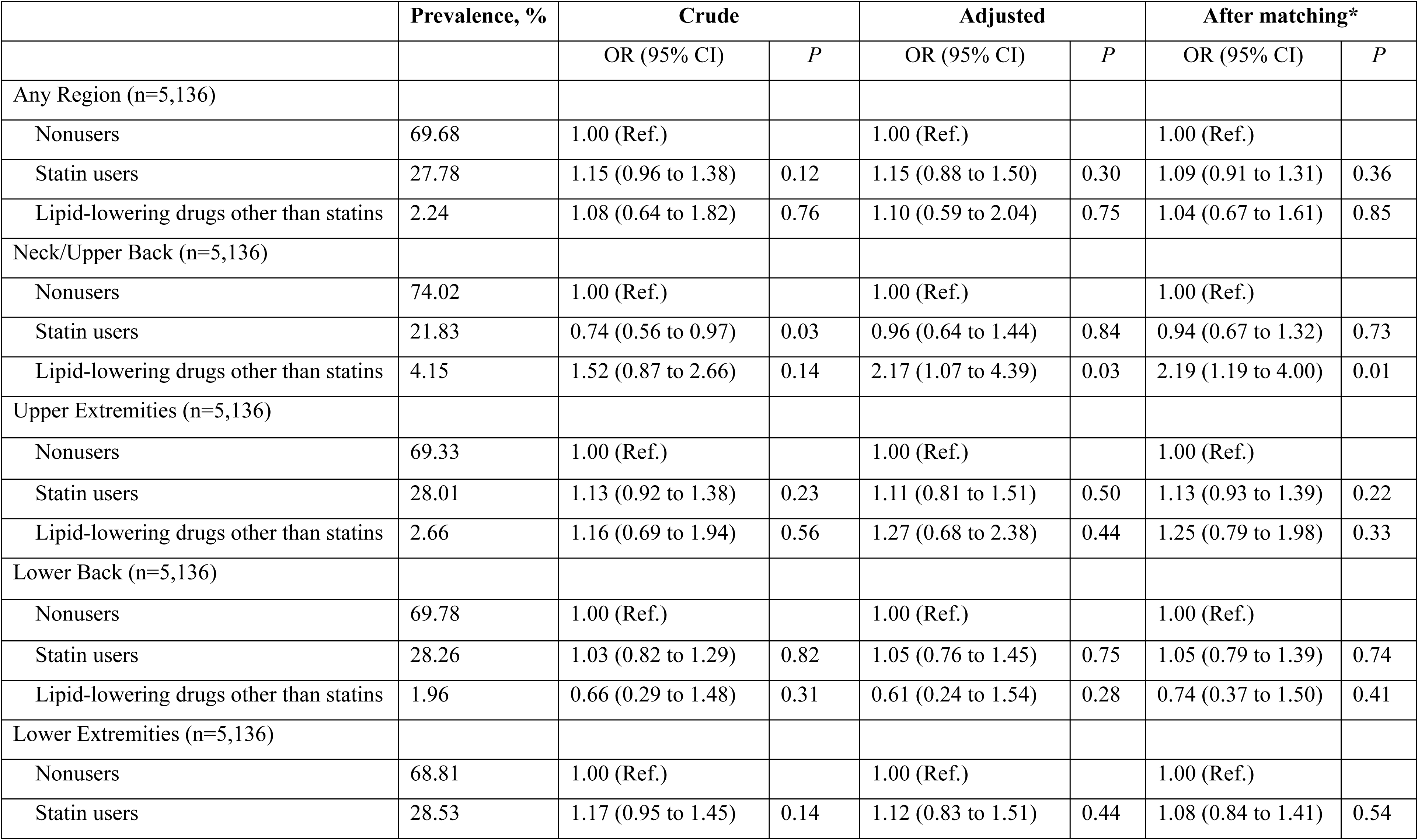
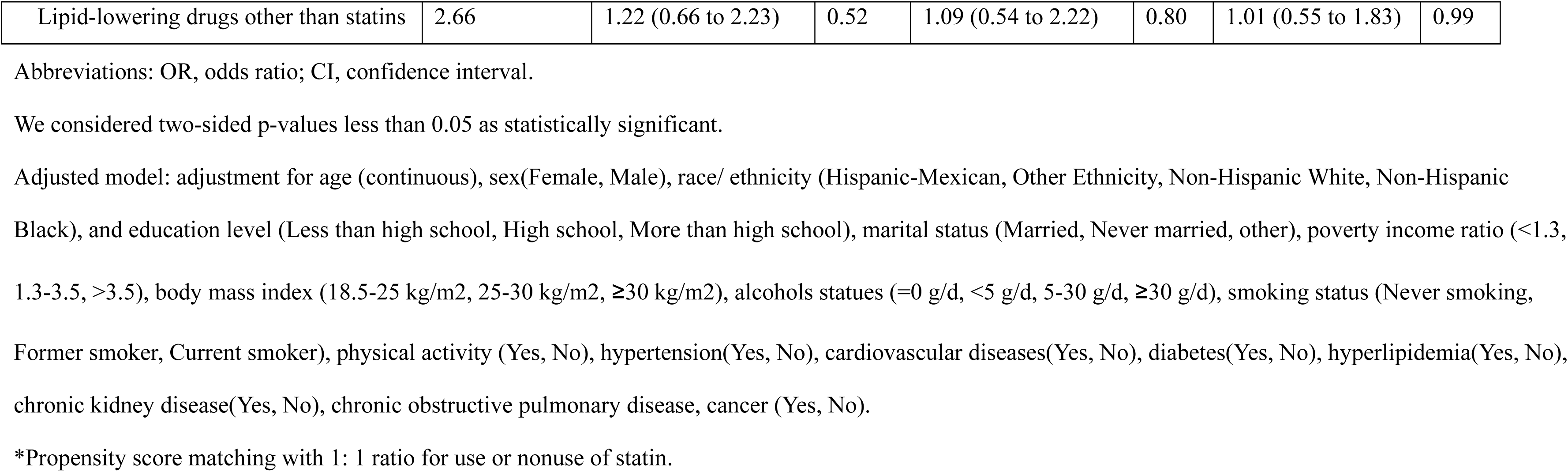
Odds ratios of musculoskeletal pain for statin users compared to nonusers.

We conducted a stepwise regression analysis to observe the association between statin use and muscle-related phenotypes and found that the associations were substantially attenuated after adjustment for age (**Table S5**).

### 3.3 MR analysis

There was moderate heterogeneity in the estimated effects of statins on the sarcopenia-related indicators appendicular lean mass, hand grip strength, and LDH (P of Cochran’s Q < 0.05) but no pleiotropy (intercept > P of 0.05) (**Table S6**). Overall IVW MR analysis (**Table 4**) revealed that compared without a statin, statin user was not associated with a higher likelihood of muscle-related phenotypes including appendicular lean mass (β=-0.060, 95% CI: -0.153 to 0.033), right-hand grip strength (β=-0.014, 95% CI: -0.039 to 0.012), left-hand grip strength (β=-0.216, 95% CI: - 0.044 to 0.001), low hand grip strength (OR=1.042, 95% CI: 0.889 to 1.221), walking pace (β=0.001, 95% CI: -0.010 to 0.011), and LDH (β=-0.080, 95% CI: -0.230 to 0.053).

**Table 4.** The causal relationship between statins and sarcopenia-related traits based on two-sample Mendelian randomization analysis.

| Outcome | SNPs | MR method | Beta/OR | 95%CI | P-value |
| --- | --- | --- | --- | --- | --- |
| **Appendicular lean mass** | 11 | IVW | -0.060 | -0.153, 0.033 | 0.208 |
|  |  | MR Egger | -0.019 | -0.191, 0.153 | 0.834 |
|  |  | Weighted median | -0.023 | -0.056, 0.010 | 0.173 |
|  | 4 | MR PRESOO | -0.016 | -0.057, 0.025 | 0.488 |
| **Hand grip strength (right)** | 11 | IVW | -0.014 | -0.039, 0.012 | 0.289 |
|  |  | MR Egger | -0.003 | -0.050, 0.043 | 0.892 |
|  |  | Weighted median | -0.010 | -0.031, 0.011 | 0.339 |
|  | 8 | MR PRESOO | 0.003 | -0.017, 0.023 | 0.776 |
| **Hand grip strength (left)** | 11 | IVW | -0.216 | -0.044, 0.001 | 0.059 |
|  |  | MR Egger | -0.004 | -0.044, 0.036 | 0.847 |
|  |  | Weighted median | -0.013 | -0.031, 0.006 | 0.171 |
|  | 10 | MR PRESOO | -0.021 | -0.041, 0.002 | 0.058 |
| **Low hand grip strength** | 11 | IVW | 1.042 | 0.889, 1.221 | 0.614 |
|  |  | MR Egger | 1.025 | 0.759, 1.384 | 0.875 |
|  |  | Weighted median | 1.028 | 0.839, 1.259 | 0.792 |
|  |  | MR PRESOO | 1.069 | 0.917, 1.247 | 0.411 |
| **Walking pace** | 11 | IVW | 0.001 | -0.010, 0.011 | 0.939 |
|  |  | MR Egger | 0.007 | -0.012, 0.026 | 0.497 |
|  |  | Weighted median | -0.002 | -0.016, 0.011 | 0.748 |
|  |  | MR PRESOO | 0.003 | -0.007, 0.012 | 0.649 |
| **Lactate dehydrogenase** | 8 | IVW | -0.080 | -0.213, 0.053 | 0.238 |
|  |  | MR Egger | 0.035 | -0.230, 0.230 | 0.804 |
|  |  | Weighted median | 0.003 | -0.082, 0.087 | 0.952 |
|  | 6 | MR PRESOO | -0.030 | -0.100, 0.040 | 0.437 |

Abbreviations: SNP, Single nucleotide polymorphism; IVW, Inverse variance weighted; MR-Egger pleiotropy test, Mendelian Randomization-Egger pleiotropy test; MR-PRESSO, MR-pleiotropy residual sum and outlier.

### 3.4 Sensitivity analysis

Sensitivity analysis revealed that among the population aged 65 and over (**Table S7**), population with cardiovascular disease (**Table S8**), population with diabetes (**Table S9**), and population with hyperlipidemia (**Table S10**), statin user was not associated with a higher likelihood of muscle-related phenotypes.

**Table S11** shows the relationships between statin use and muscle-related phenotypes in the redefined hyperlipidemic population. Propensity score matching revealed that statin user was not associated with a higher likelihood of muscle-related phenotypes including ASM/BMI (OR=1.25, 95% CI: 0.99 to 1.59), ASM/Ht^2^ (OR=0.94, 95% CI: 0.66 to 1.33), maximum HGS (β=- 0.45, 95% CI: -1.04 to 0.14), relative HGS(β=-0.03, 95% CI: -0.07 to 0.01), combined HGS (β=-2.33, 95% CI: - 5.47 to 0.81), ASM/Ht^2^ and body fat percentage defined as sarcopenic obesity (OR=1.23, 95% CI: 0.96 to 1.57), ASM and BMI defined as sarcopenic obesity (OR=0.85, 95% CI: 0.53 to 1.34), LDH (β=- 0.62, 95% CI: -4.69 to 3.46), any region pain (OR=1.10, 95% CI: 0.89 to 1.35), neck/upper back pain (OR=0.89, 95% CI: 0.61 to 1.30).

## 4. Discussion

In this nationally representative cross-sectional study with 22,549 patients with a history of cardiovascular disease, diabetes, or hyperlipidemia, 8,481 (37.6%) participants used statin. We found that in the unadjusted model, compared with adults not using any lipid-lowering drugs, statin use was associated with a higher likelihood of sarcopenia, sarcopenic obesity, lower HGS, and combined HGS. However, after multivariable adjustment or propensity score match, the independent associations of statin use with sarcopenia, sarcopenic obesity, HGS, LDH, and musculoskeletal pain became nonsignificant. Stepwise regression suggested that age was the predominant confounding factor for these associations. MR analysis also showed no significant causality between statin use and skeletal muscle-related phenotypes including appendicular lean mass, hand grip strength (left and right), low hand grip strength, walking pace, or LDH.

Our results are inconsistent with previous studies, in which we found no association between statin use and skeletal muscle-related phenotypes. Chronic heart failure resulting from various cardiovascular diseases may contribute to secondary sarcopenia related to disease in patients with cardiovascular disease[32]. Notably, sarcopenia is associated with worse quality of life and increased mortality in patients with CVD[32]. A meta-analysis found significant differences in myopathic symptoms and muscle pain between patients on low- and high-intensity statin therapy. However, only 57% of myopathy symptoms and 16% of muscle pain were dose-related[33]. In a large observational study of 46,249 participants, statin users exhibited higher rates of arthropathy, musculoskeletal injuries, and pain compared to controls matched for age, sex, and comorbidities[34]. While this study found no association between statin use and increased likelihood of musculoskeletal pain among individuals with cardiovascular disease, diabetes, or hyperlipidemia, another observational study reported a significantly higher incidence of musculoskeletal pain in various regions, including the lower back and lower extremities, among statin users without arthritis[13]. The study revealed that statin user was not associated with a higher likelihood of maximum HGS, relative HGS, or combined HGS, and other observational results conflicted. For instance, findings from the Hertfordshire cohort study indicated that calcium channel blockers and fibrates were linked to reduced grip strength in women, whereas statins showed no such association[17].

There is no evidence of an increased likelihood of muscle-related phenotypes in the population receiving statins. Data from RCTs on the efficacy of statins in patients aged 65 years and older showed that[35] although older adults are at a higher risk for cardiovascular disease and are most likely to benefit from lipid-lowering therapy, they are the group least likely to receive statins[36]. This suggests that statins are still underutilized, with patients >65 years less likely to receive a statin prescription than younger patients[37]. This phenomenon primarily stems from concerns among older adults regarding myopathy as an adverse reaction linked to statin therapy. However, this cross-sectional study revealed statin user was not associated with a higher likelihood of muscle-related phenotypes including sarcopenia, HGS, or LDH in the population over 65 years of age, indicating that in terms of adverse muscle effects, statins may play a role in cardiovascular disease, diabetes, or relative safety in patients with hyperlipidemia.

Our results found that the association between statin use and muscle-related phenotypes was significantly attenuated after adjustment for age. Muscle complaints are commonly reported by older adults and can stem from various causes, including sarcopenia, increased physical activity, diseases that predispose to or exacerbate muscle issues, and medications known for their myotoxic effects. This misinterpretation could lead to a significant number of older adults avoiding statin therapy, thereby missing out on potential cardiovascular benefits and experiencing more unintended events. One randomized controlled trial revealed that 90% of the symptoms experienced while taking statins also occurred while taking a placebo[38]. Furthermore, a recent systematic review revealed that between 38% and 78% of individuals reported statin-related muscle symptoms[39]. A recent meta-analysis proposed a “nocebo effect”, which occurs when subjective adverse effects, such as aches and pains, result from patients’ anticipation of harm from statin therapy due to their awareness of and concern about potential side effects. A meta-analysis of randomized clinical trials on statins indicated comparable rates of adverse events between participants assigned to statins or placebo, suggesting no notable increase in adverse event rates associated with statin use[39]. However, many researchers argue that because trial populations are often healthier than real-world clinical populations, observational studies can more accurately estimate the frequency of adverse events in clinical practice.

The findings of our study demonstrate the long-term safety of statin use, which can enhance patient compliance and address the concerns of both users and healthcare providers. Concurrently, the aging of the population will undoubtedly increase the incidence of musculoskeletal disorders closely associated with aging, including sarcopenia, osteoporosis, and osteopenia[40], therefore muscle-related symptoms should be examined through the lens of the aging process. Our study provides a convincing supplement to the current small sample RCTs, which are insufficient in terms of generalizability and outreach.

### 4.1 Limitations and Strengths

The strengths of the present study include its large and representative sample, facilitating adjustment for numerous confounding factors and thereby enhancing the reliability of the results. Furthermore, propensity score matching was employed to balance confounding factors related to key medications, thereby reducing potential drug interactions. The main strength of this study lies in combining observational data from the National Nutrition Health and Nutrition Examination Survey 1999–2020 with the MR approach.

Our study has several limitations. First, the cross-sectional design prevents us from establishing causality or accounting for individuals who may have initiated or ceased statin use due to skeletal muscle adverse events, which were assessed using MR analysis to explore causal relationships. Second, we assessed statin use only for the month before the interview, so we could not determine the duration of statin use or whether non-users had recently stopped taking statins. Additionally, our study faced limitations in obtaining enough statin users to evaluate the effects of various types of statins on outcomes. We also lacked dosage information to investigate potential associations with high statin doses, which contribute to statin-induced muscle-related side effects[13]. Future studies should gather comprehensive medication use histories, including dosage details from prescription labels, frequency, duration of use, and discontinuation patterns. This would enhance the robustness of the study findings. Third, due to the absence of creatine kinase level data, we were unable to investigate the association between statin or other lipid-lowering drug use and creatine kinase levels. These data, combined with longitudinal follow-up, could provide further insights into the musculoskeletal side effects of statins, despite this study utilizing LDH as a muscle damage marker. Moreover, our study sample was representative of the U.S. population, primarily comprising self-reported non-Hispanic White participants. However, data on individuals of Asian descent were not captured. This detail is crucial as research suggests that individuals of Asian descent may exhibit higher susceptibility to statin-induced musculoskeletal side effects compared to other demographic groups. Fourth, this study only focused on skeletal muscle-related phenotypes. Other side effects such as diabetes, cognition, and liver damage require special research. Furthermore, data exclude hospitalized patients, who typically exhibit poorer health statuses, multiple comorbidities, and potentially heightened susceptibility to adverse drug reactions. Last, in this observational study, despite efforts to control for numerous critical confounding factors, residual and unmeasured confounders may have constrained the findings.

### 4.2 Conclusions

Our epidemiological and MR analyses did not support the causality between statin use and skeletal muscle-related phenotypes. A higher likelihood of skeletal muscle-related adverse phenotypes in statin users may be attributed to age. Future studies should further explore the biological factors that may affect the statin-related muscle phenotype. These findings will provide evidence for the drug safety of statin therapy for the secondary prevention of cardiovascular disease, diabetes, or hyperlipidemia.

## Data Availability

The datasets supporting the conclusions of this article are available in the National Health and Nutrition Examination Survey repository, unique persistent identifier and hyperlink to the dataset in https://wwwn.cdc.gov/nchs/nhanes/default.aspx.

## Ethical Standards Disclosure

This study was conducted according to the guidelines laid down in the Declaration of Helsinki and all procedures involving research study participants were approved by the [NCHS]. Written informed consent was obtained from all subjects.

## Consent for publication

Not applicable.

## Authors contributions

FT, ZC, and SW conceived and designed the study. FT, HQ, YL, ZH, and BY organized all data. FT, ZC, and SW analyzed and visualized the results. FT, YS, and SW contributed to the manuscript. Reviewed and edited the manuscript, YZ and SF take responsibility for the integrity and accuracy of this analysis. All authors reviewed and edited the manuscript.

## Acknowledgments

The authors thank all the participants and staff in the National Health and Nutrition Examination Survey for their substantial contributions.

## Supplemental tables

**Table S1.**
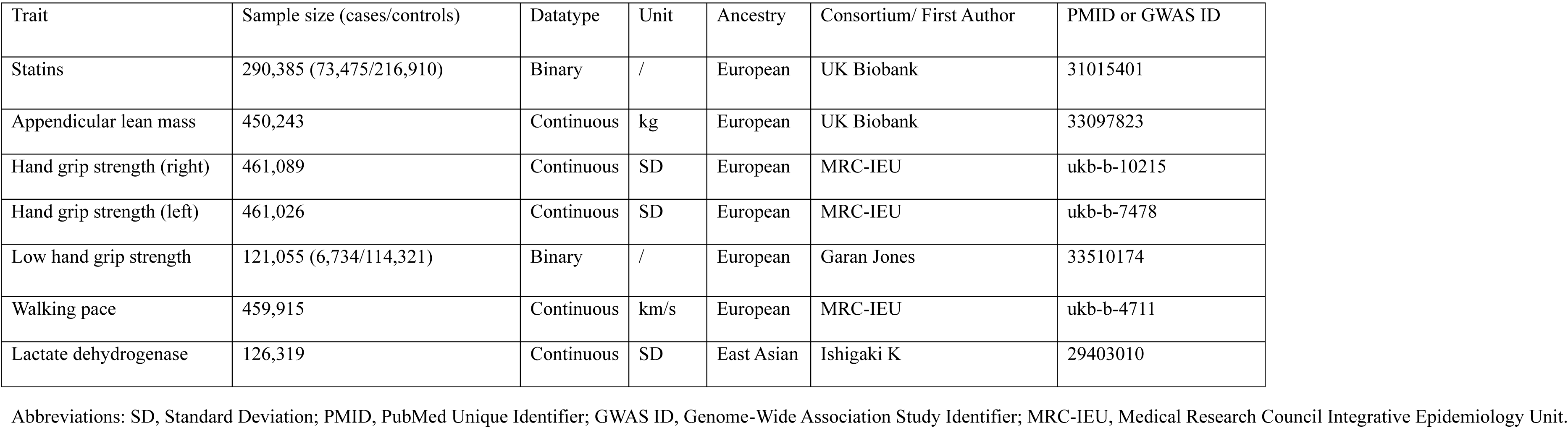
Details of studies and datasets used in the study.

| Trait | Sample size (cases/controls) | Datatype | Unit | Ancestry | Consortium/ First Author | PMID or GWAS ID |
| --- | --- | --- | --- | --- | --- | --- |
| Statins | 290,385 (73,475/216,910) | Binary | / | European | UK Biobank | 31015401 |
| Appendicular lean mass | 450,243 | Continuous | kg | European | UK Biobank | 33097823 |
| Hand grip strength (right) | 461,089 | Continuous | SD | European | MRC-IEU | ukb-b-10215 |
| Hand grip strength (left) | 461,026 | Continuous | SD | European | MRC-IEU | ukb-b-7478 |
| Low hand grip strength | 121,055 (6,734/114,321) | Binary | / | European | Garan Jones | 33510174 |
| Walking pace | 459,915 | Continuous | km/s | European | MRC-IEU | ukb-b-4711 |
| Lactate dehydrogenase | 126,319 | Continuous | SD | East Asian | Ishigaki K | 29403010 |

Abbreviations: SD, Standard Deviation; PMID, PubMed Unique Identifier; GWAS ID, Genome-Wide Association Study Identifier; MRC-IEU, Medical Research Council Integrative Epidemiology Unit.

**Table S2.**
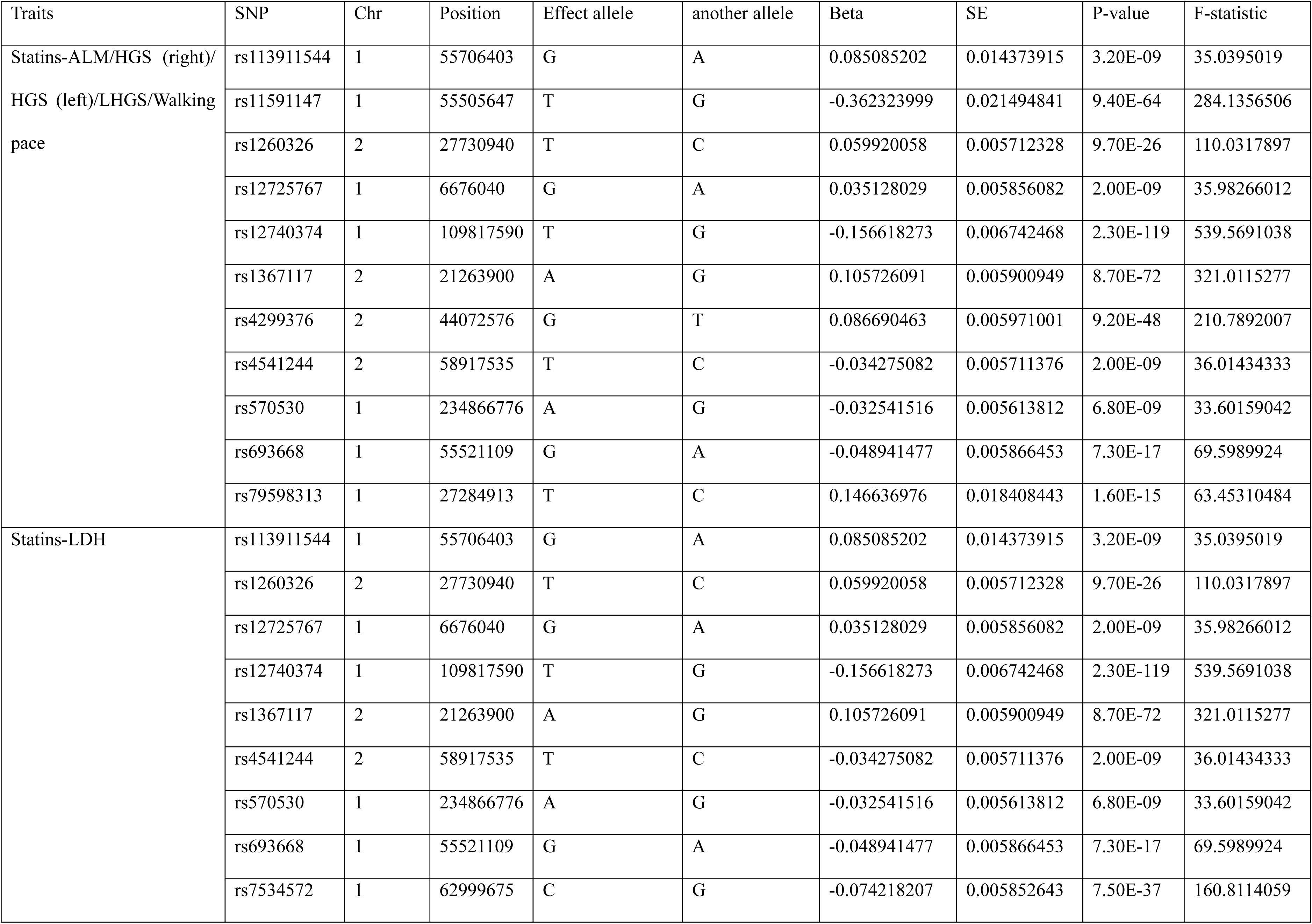
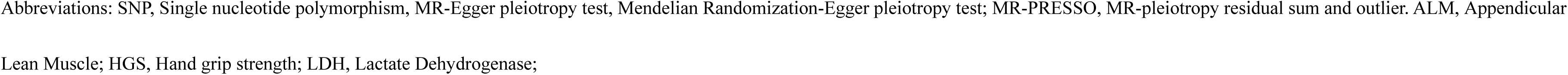
SNPs are used as instrumental variables for Statins (P-value < 5×10^-8^)

**Table S3.**
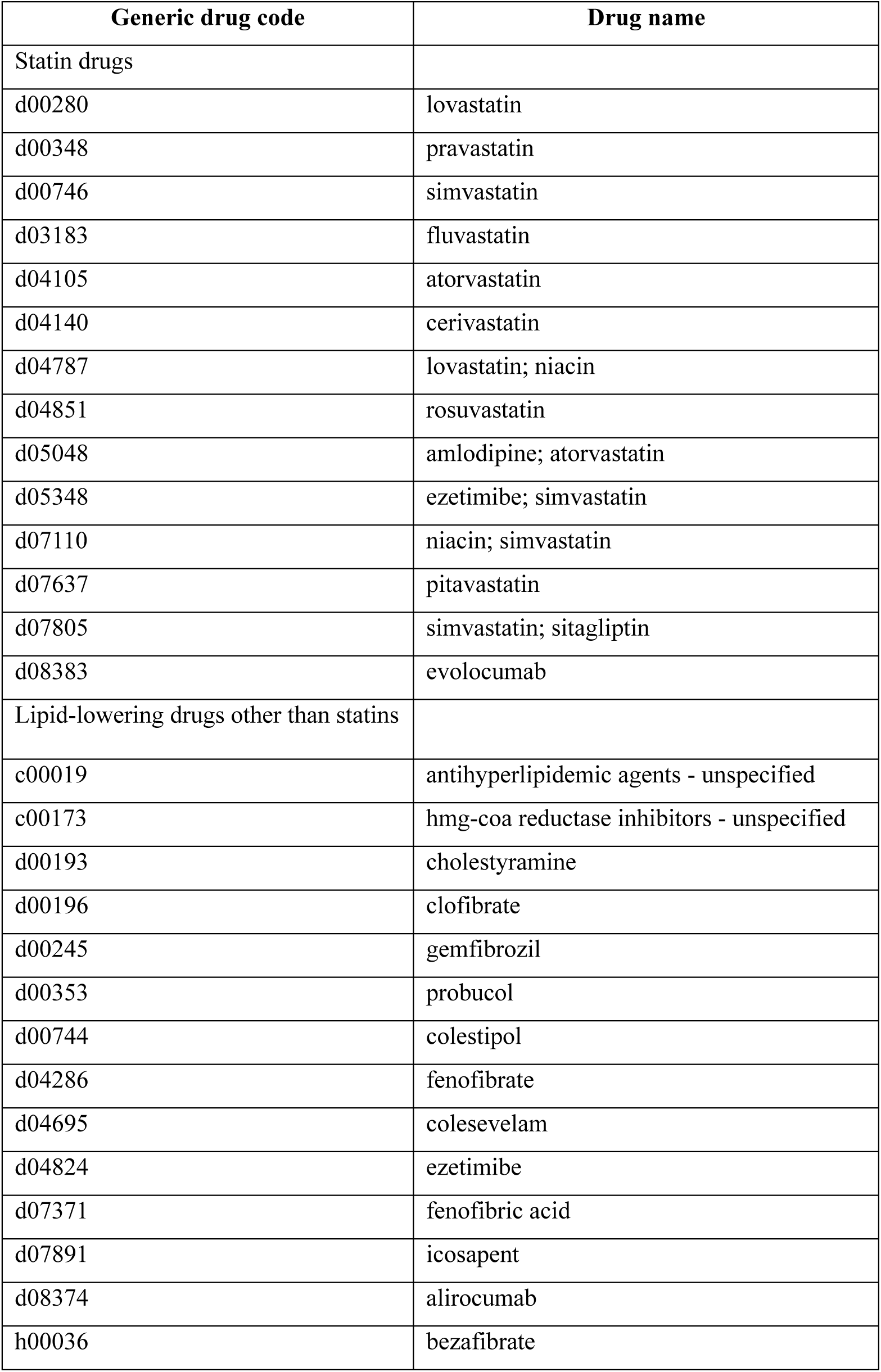
Definition of statins and lipid-lowering drugs.

**Table S4.**
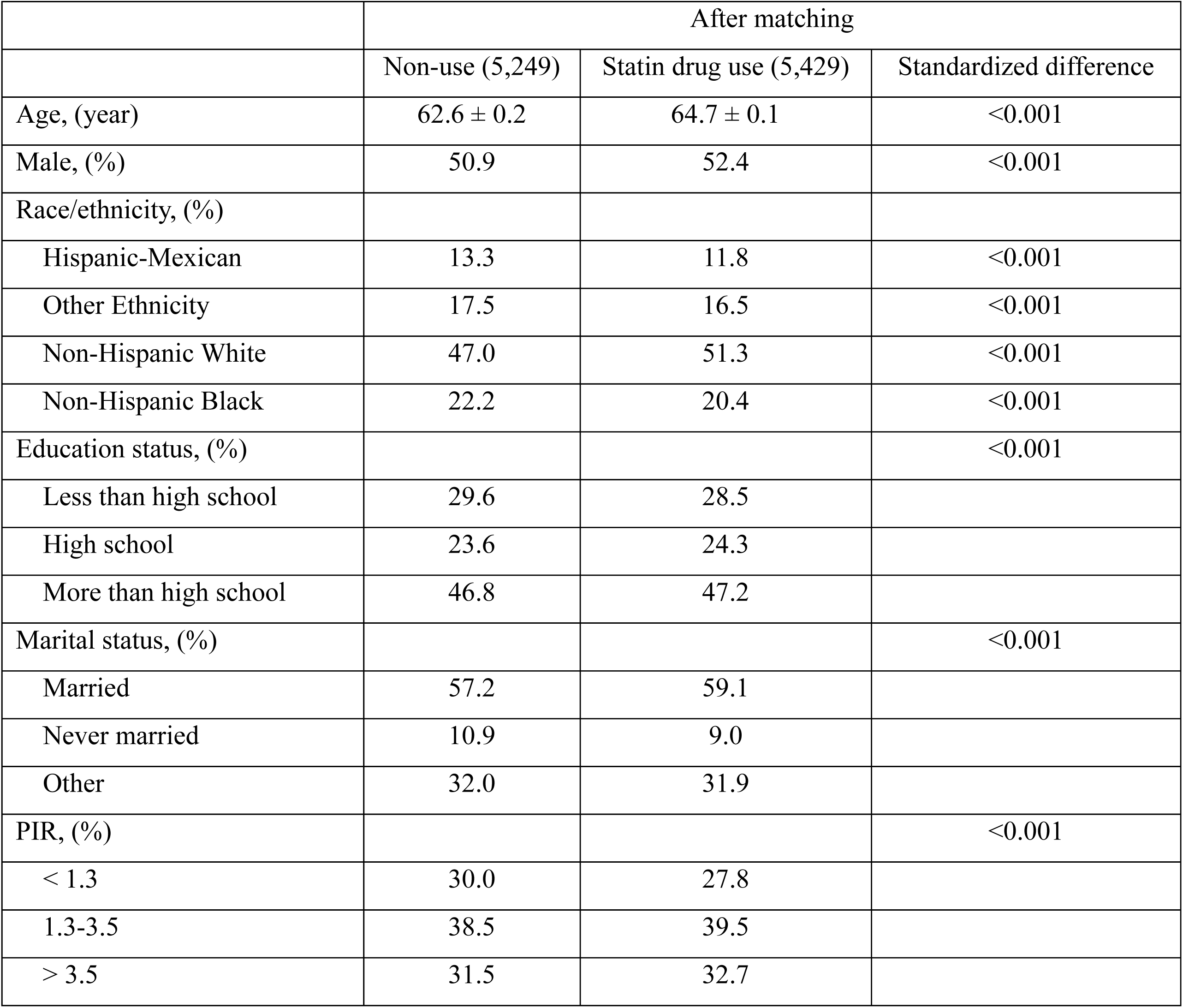
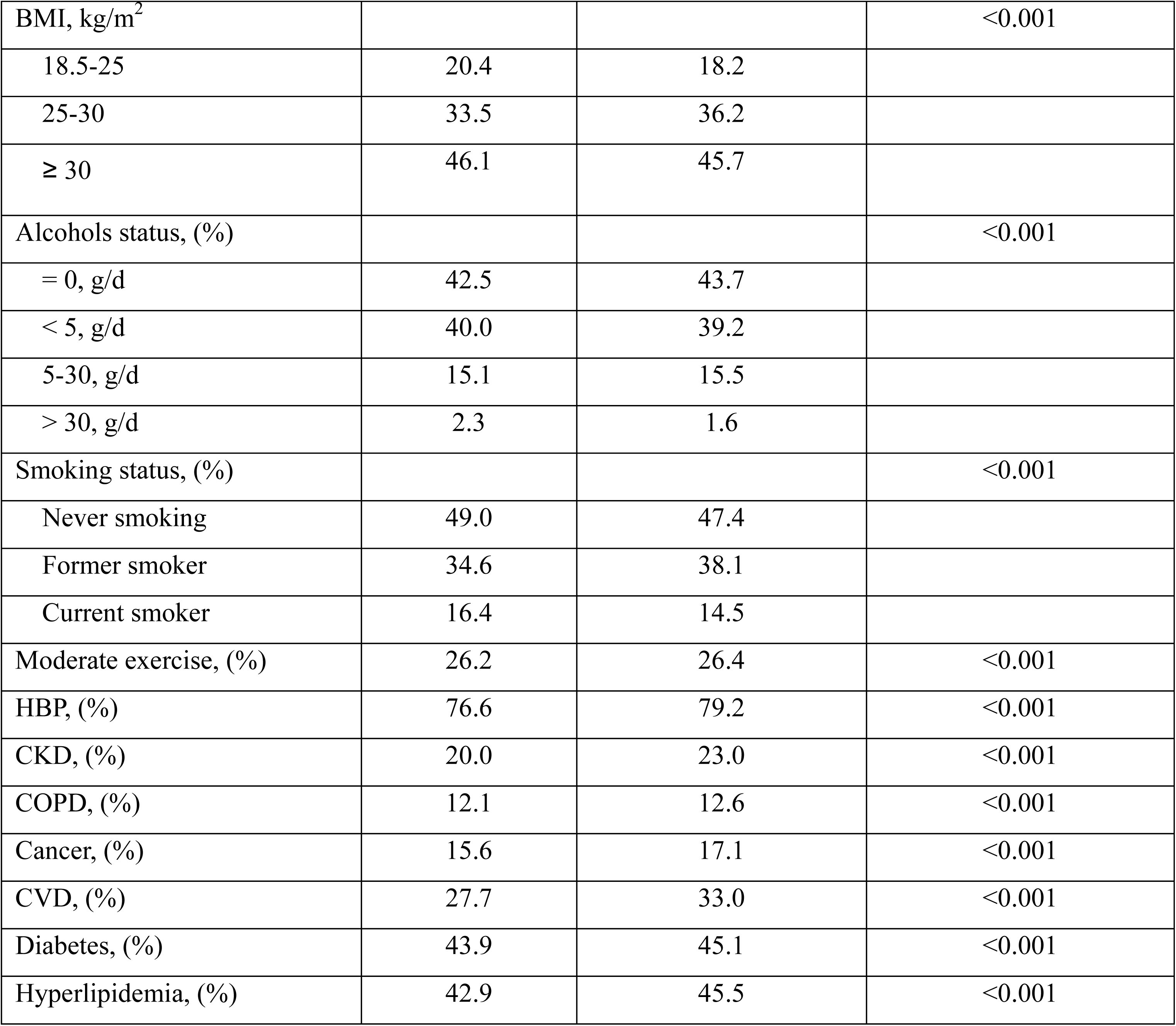
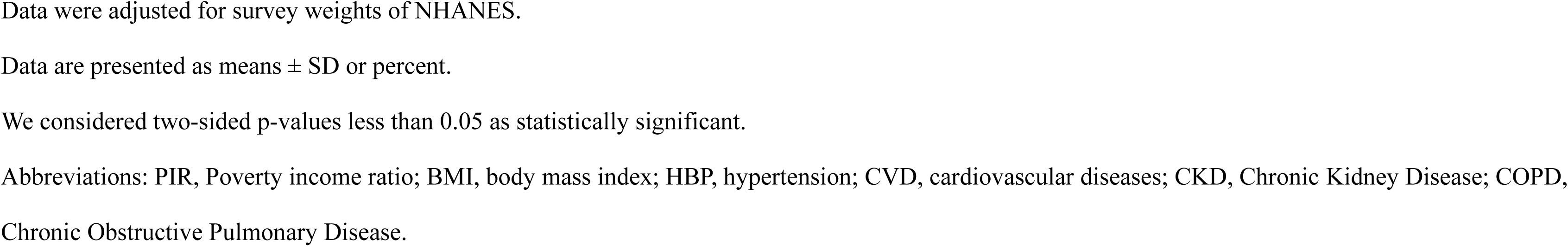
Characteristics of cardiovascular disease, diabetes, or hyperlipidemia patients with Statin use or nonuse after matching.

**Table S5.**
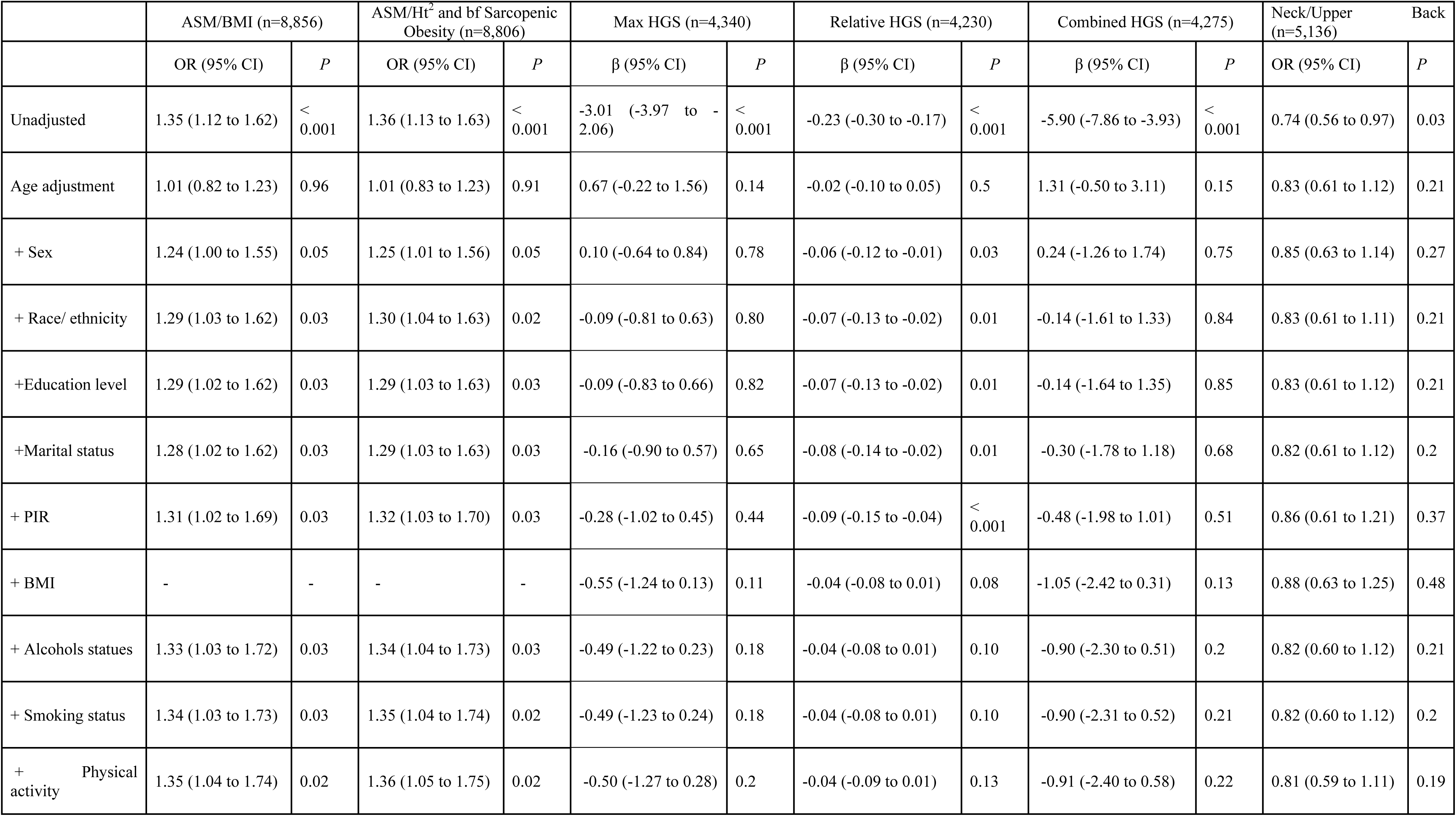
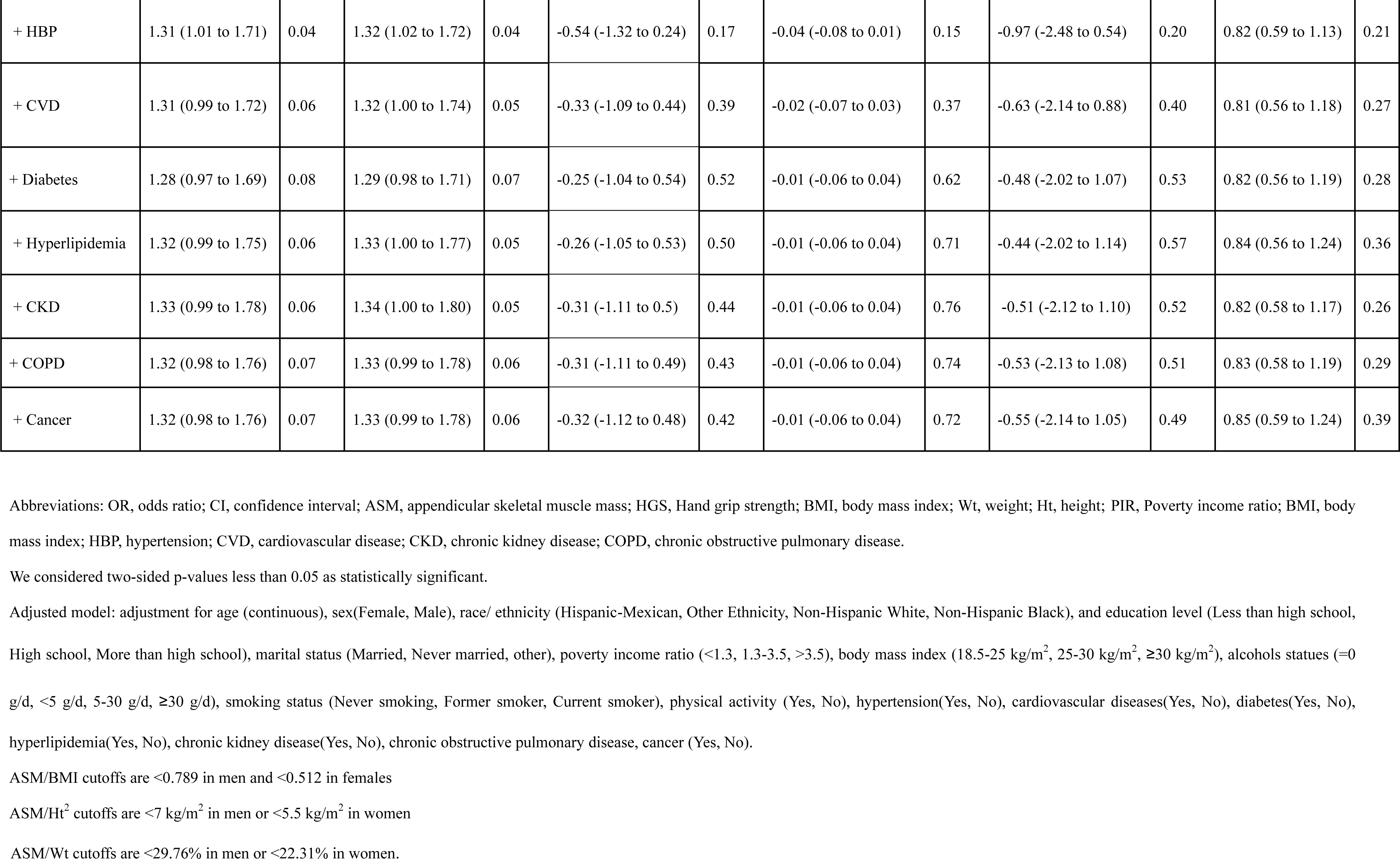
The association between statin users and skeletal muscle-related phenotypes compared with nonusers by stepwise regression analysis.

**Table S6.** The results of sensitivity analysis for two-sample Mendelian randomization analysis of the causal relationships between Statins and sarcopenia-related traits.

| Outcome | No. of IVs | Heterogeneity |  | MR-Egger pleiotropy test |  |  | MR-PRESSO |
| --- | --- | --- | --- | --- | --- | --- | --- |
|  |  | Cochran's *Q* | *P*-value | intercept | SE | *P*-value | Global Test *P*-value |
| Appendicular lean mass | 11 | 346.6756844 | 2.02E-68 | -0.0042237 | 0.00746 | 0.585 | <0.001 |
| Hand grip strength (right) | 11 | 40.71459187 | 1.27E-05 | -0.0010619 | 0.00202 | 0.612 | <0.001 |
| Hand grip strength (left) | 11 | 32.39685682 | <0.001 | -0.0018087 | 0.00172 | 0.321 | <0.001 |
| Low hand grip strength | 11 | 10.56373443 | 0.393 | 0.00164006 | 0.01293 | 0.902 | 0.463 |
| Walking pace | 8 | 8.438255201 | 0.586 | -0.0006592 | 0.00082 | 0.443 | 0.622 |
| Lactate dehydrogenase | 11 | 38.72767287 | 2.20E-06 | -0.0082974 | 0.00844 | 0.363 | <0.001 |

Abbreviations: MR-Egger pleiotropy test, Mendelian Randomization-Egger pleiotropy test; MR-PRESSO, MR-pleiotropy residual sum and outlier.

**Table S7.**
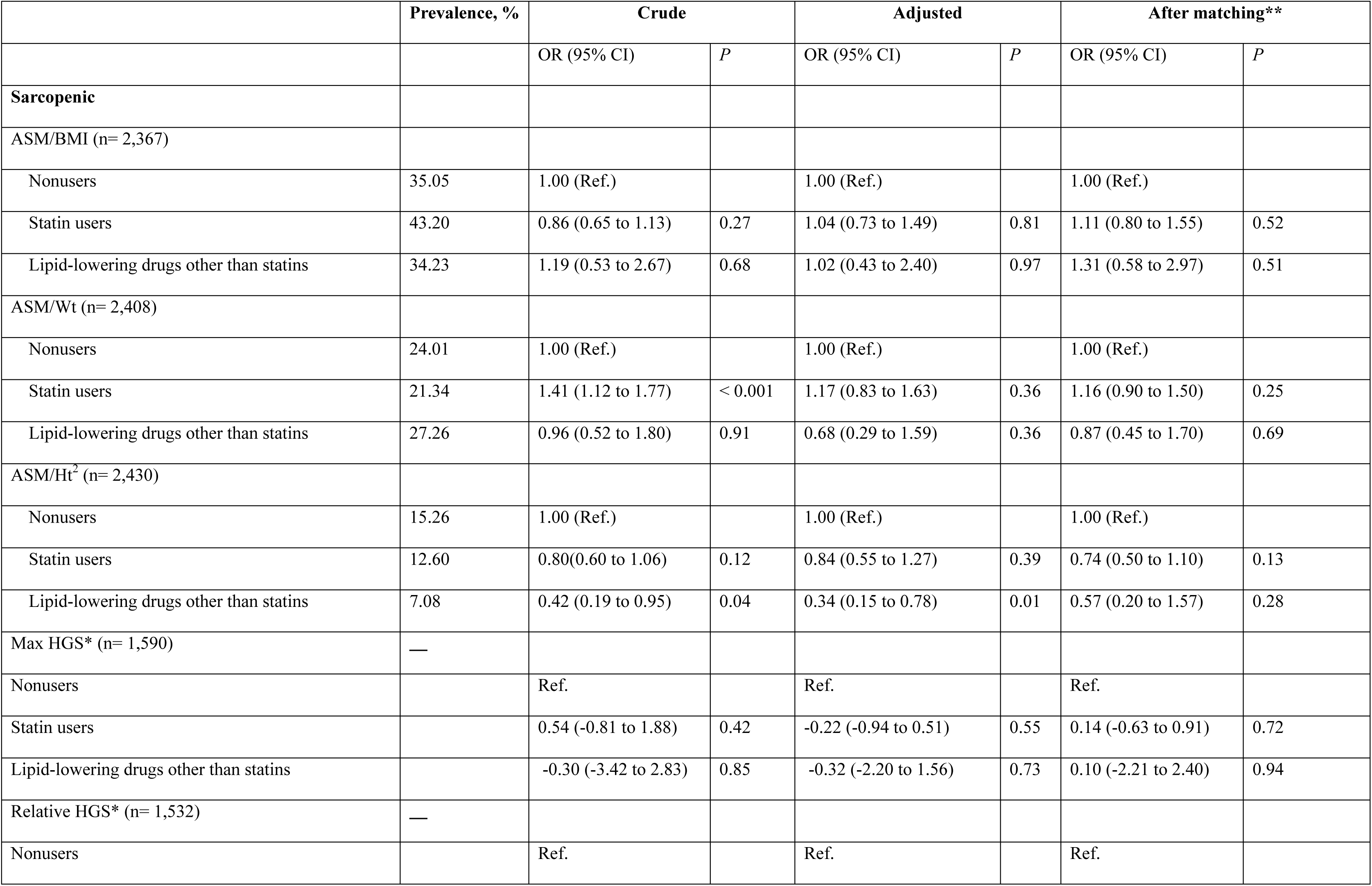
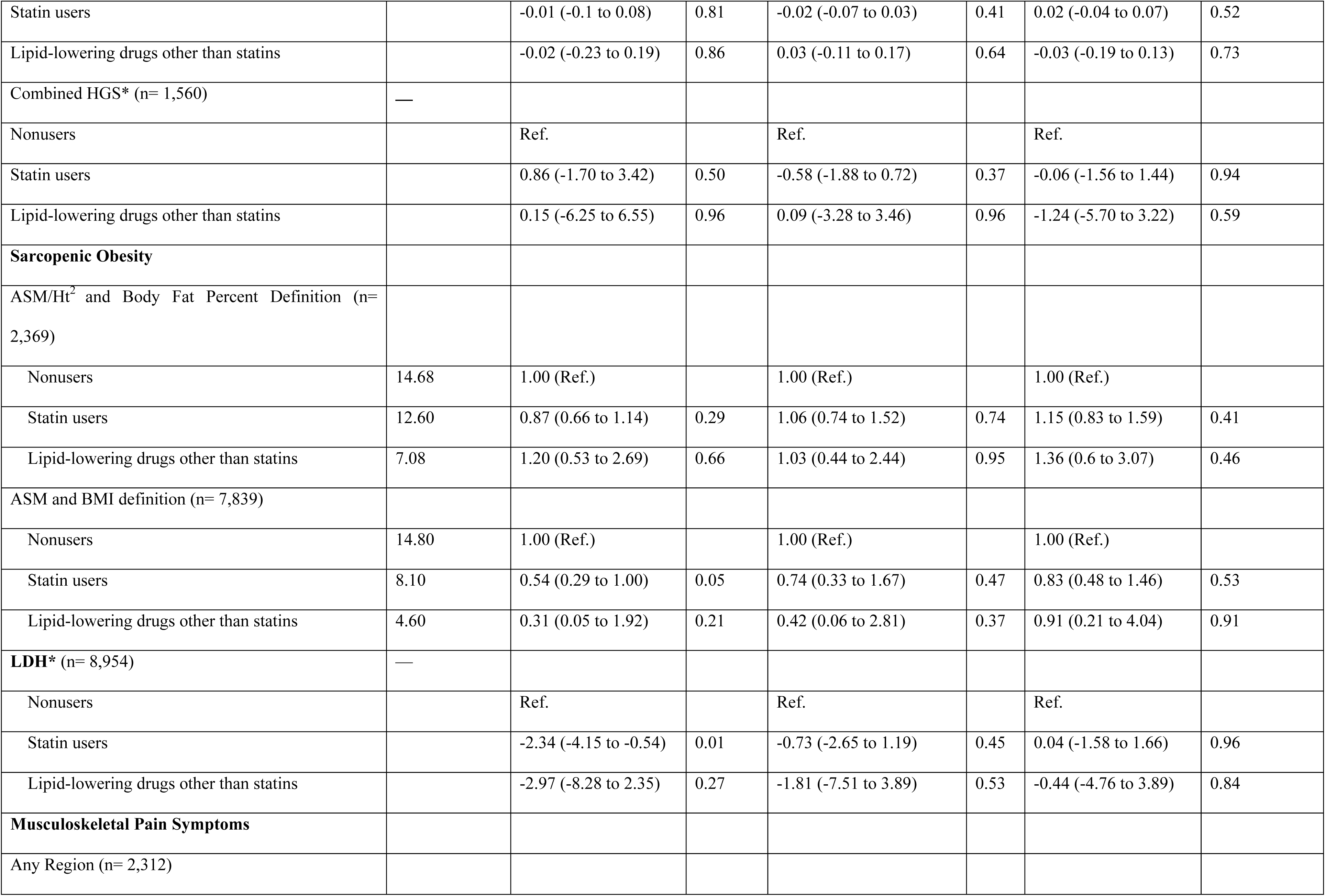
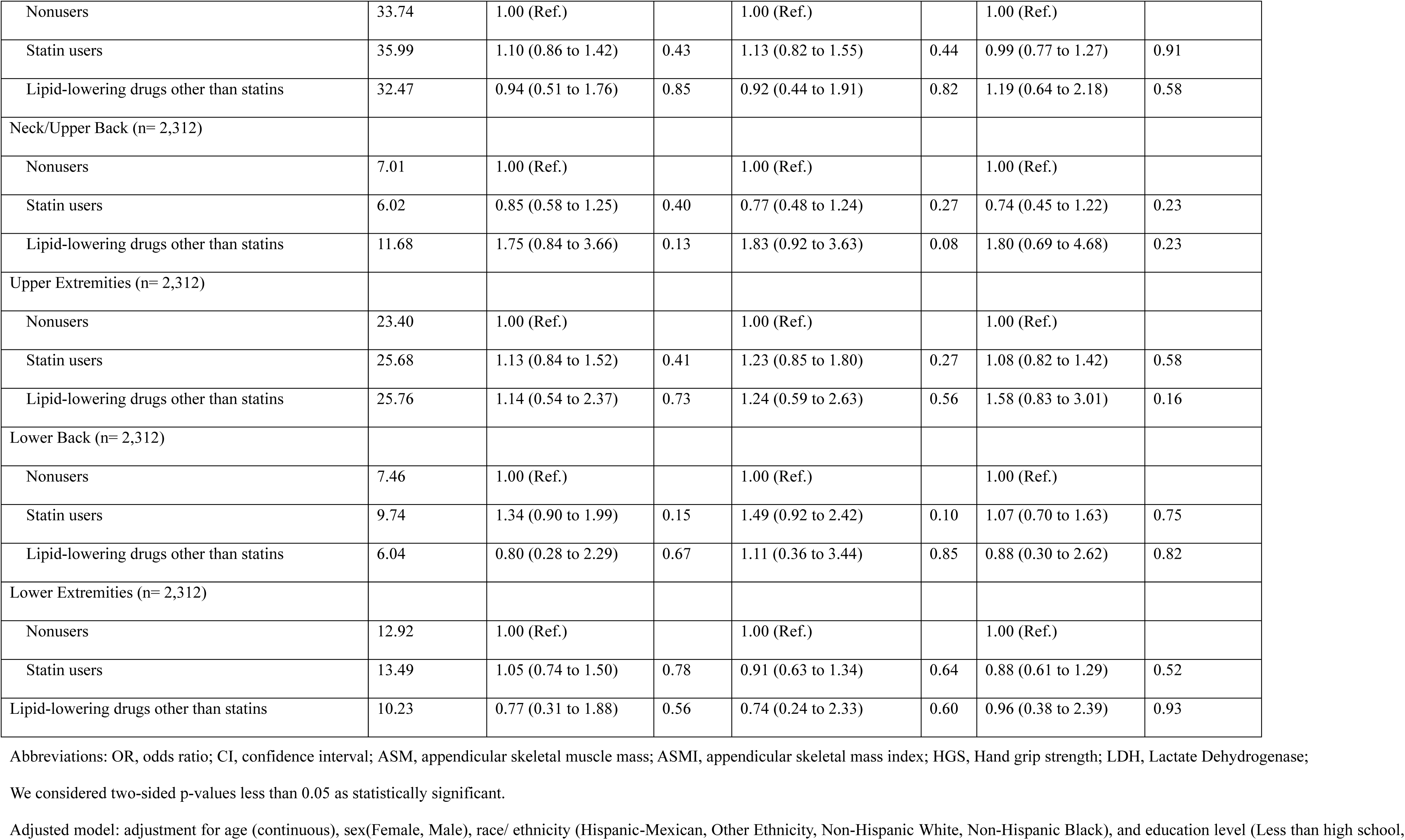
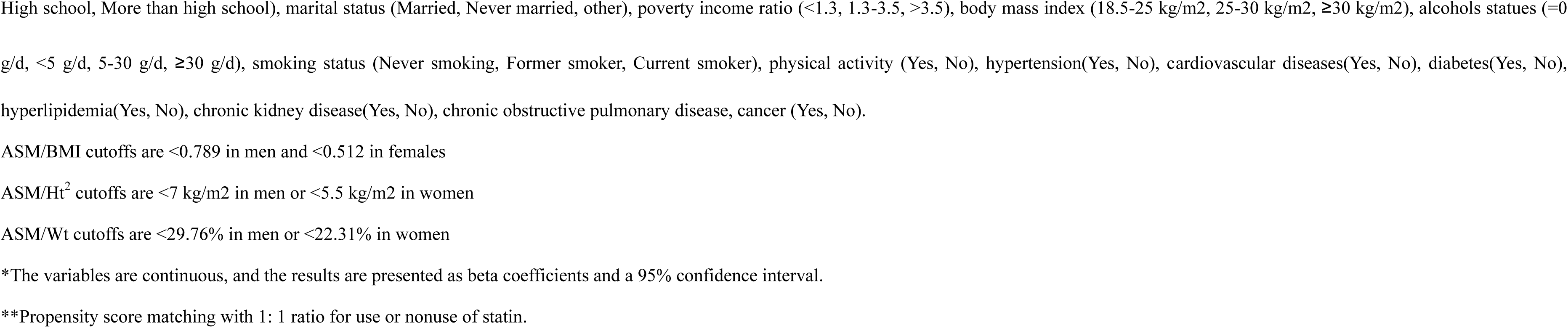
The association between statin users or lipid-lowering users and skeletal muscle-related phenotypes compared with nonusers among the population over 65 years old.

**Table S8.**
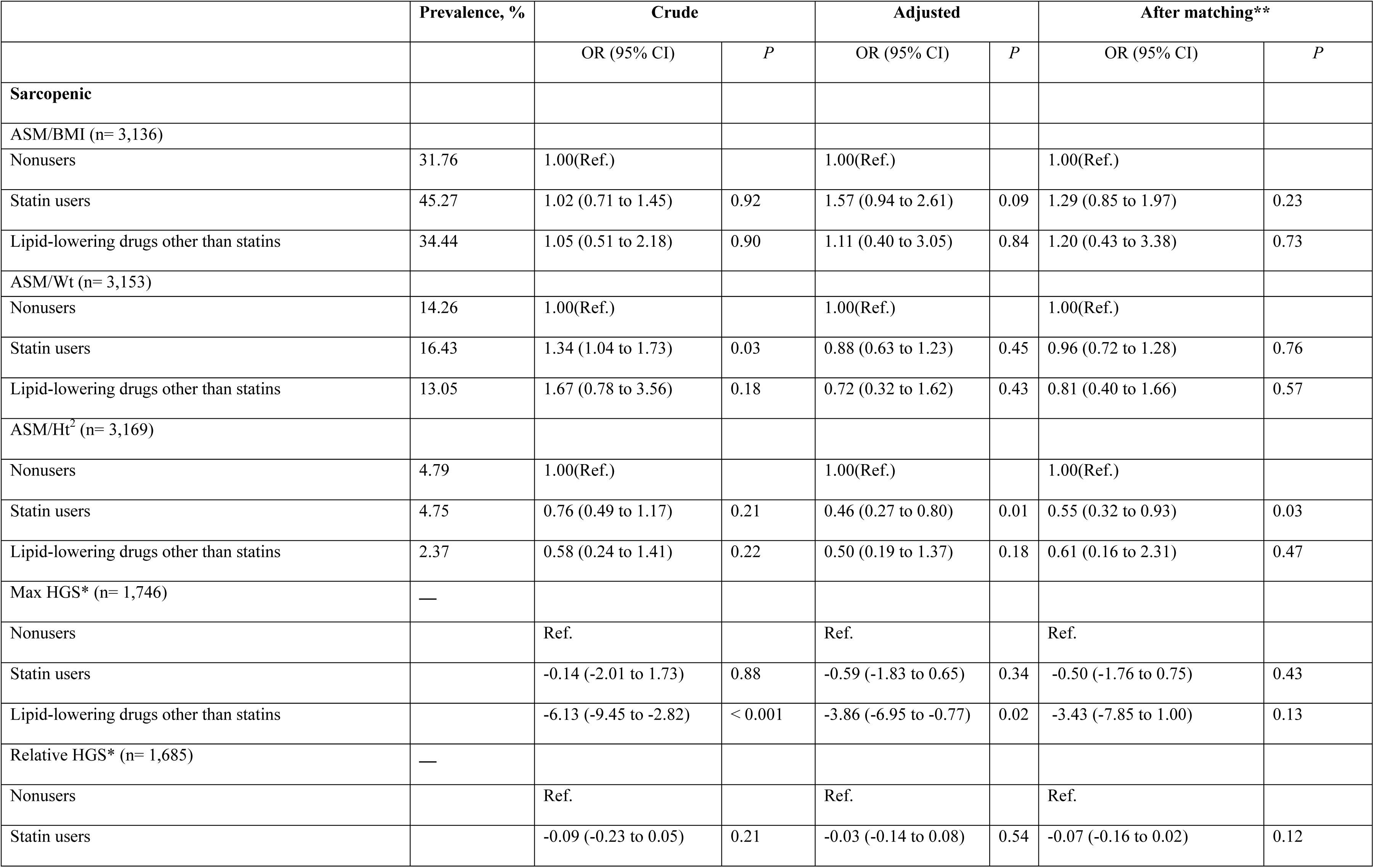
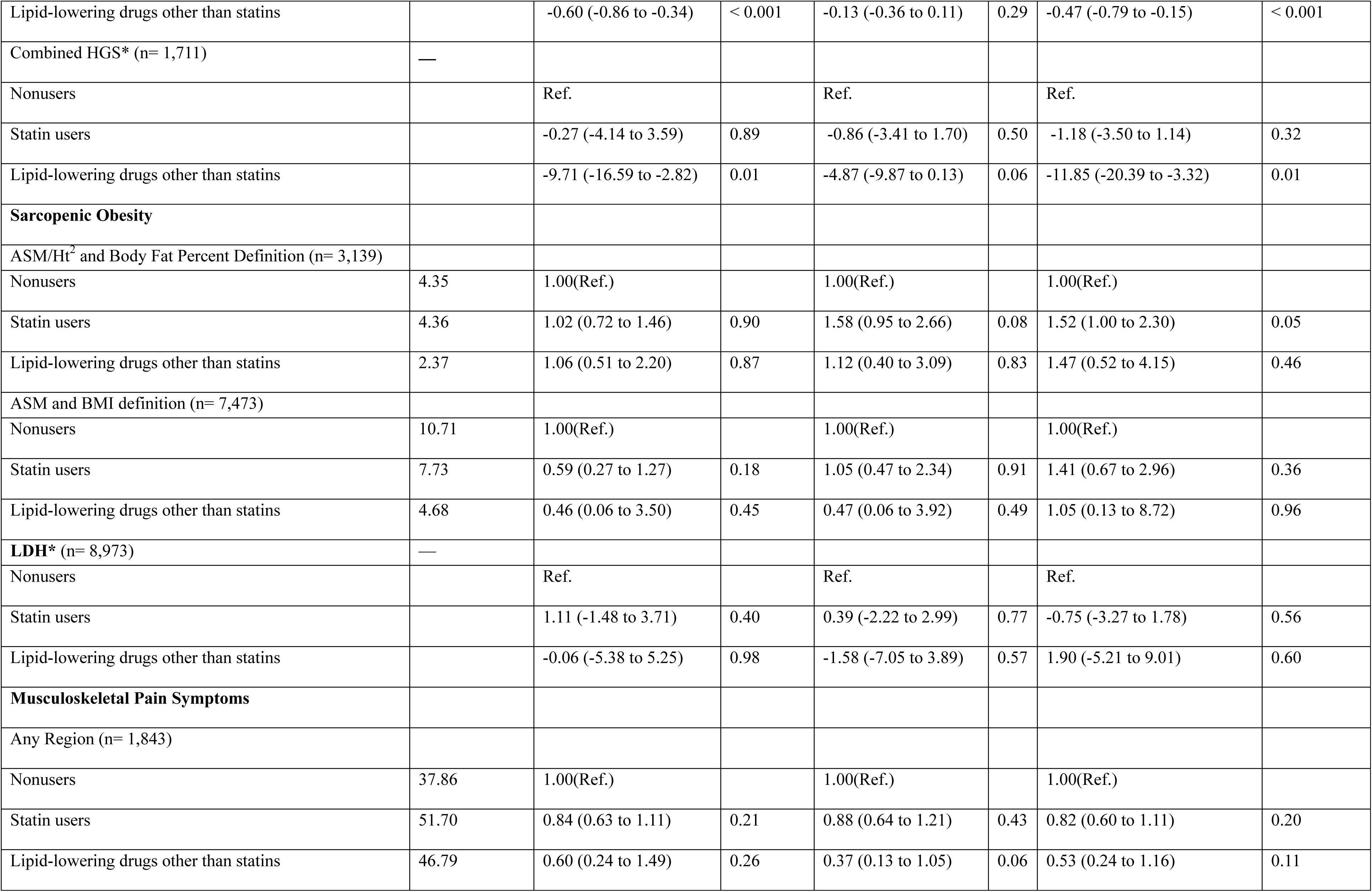
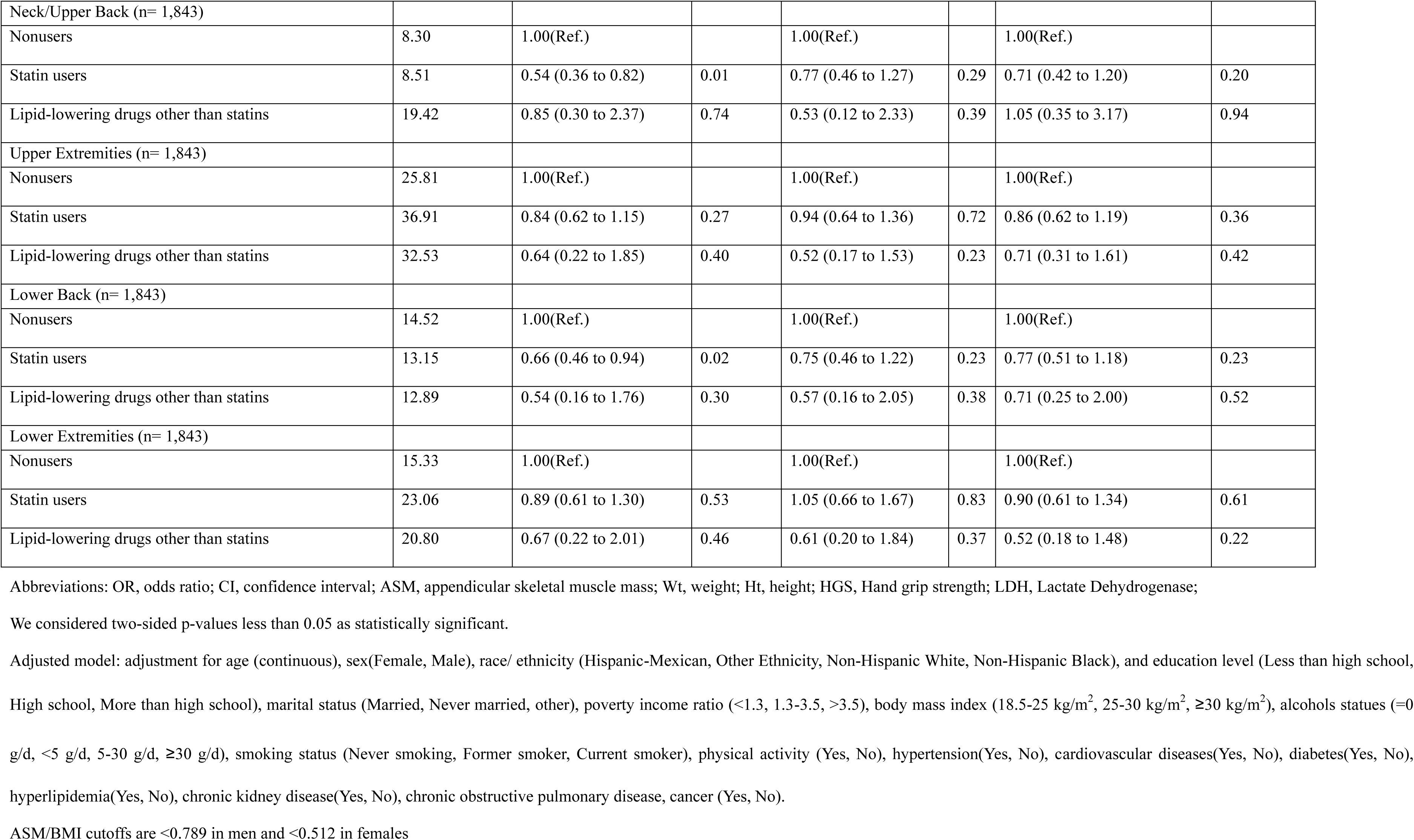
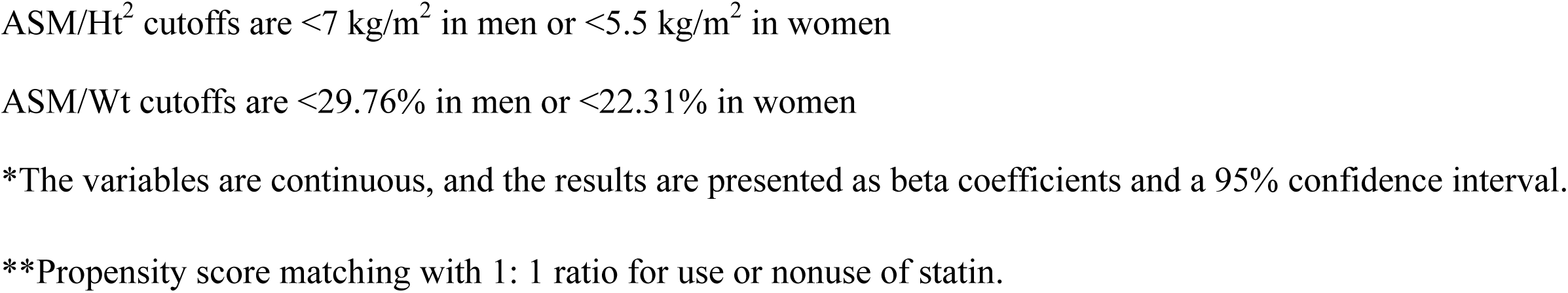
The association between statin users or lipid-lowering users and skeletal muscle-related phenotypes compared with nonusers among participants with diabetes.

**Table S9.**
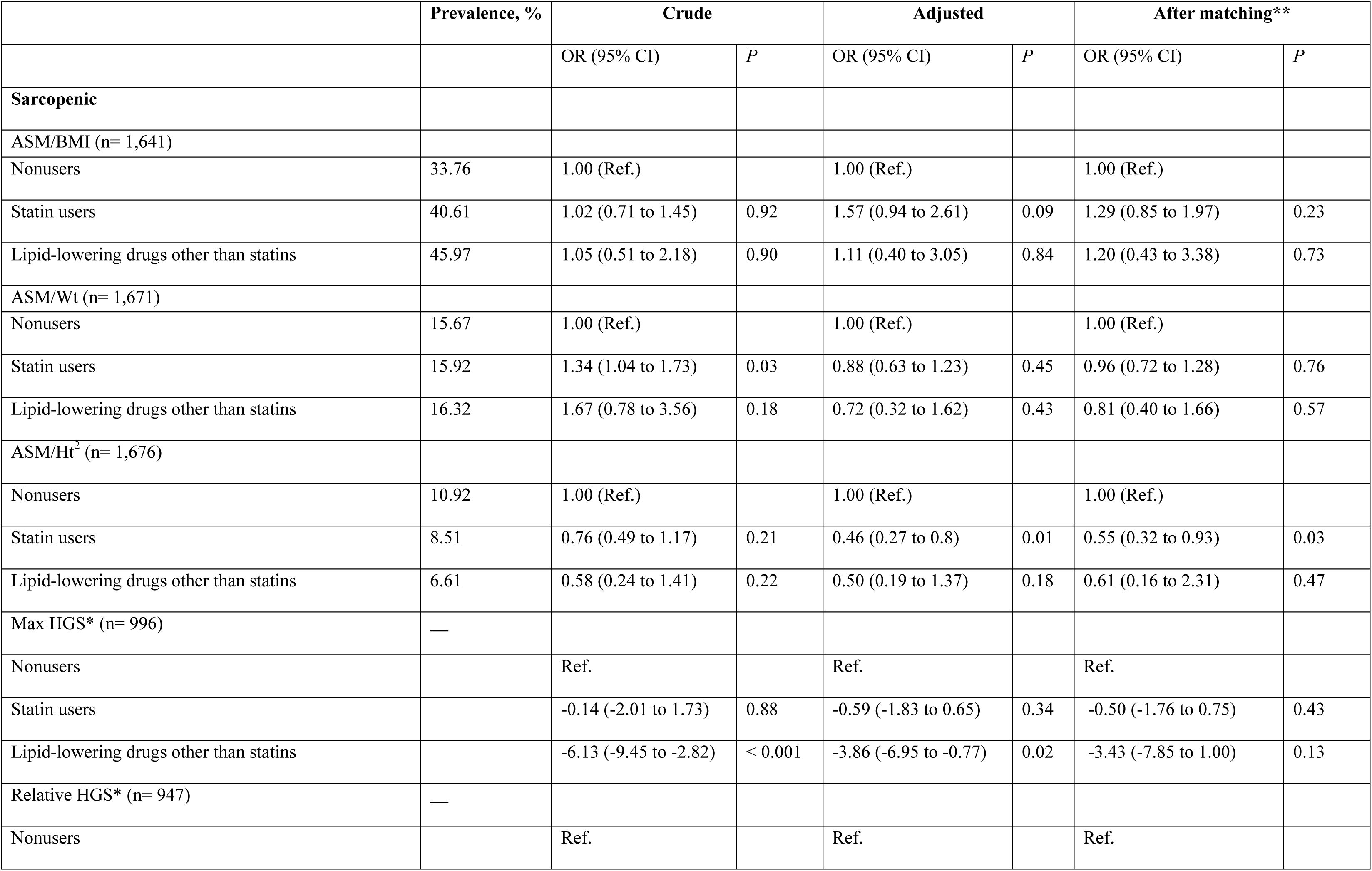
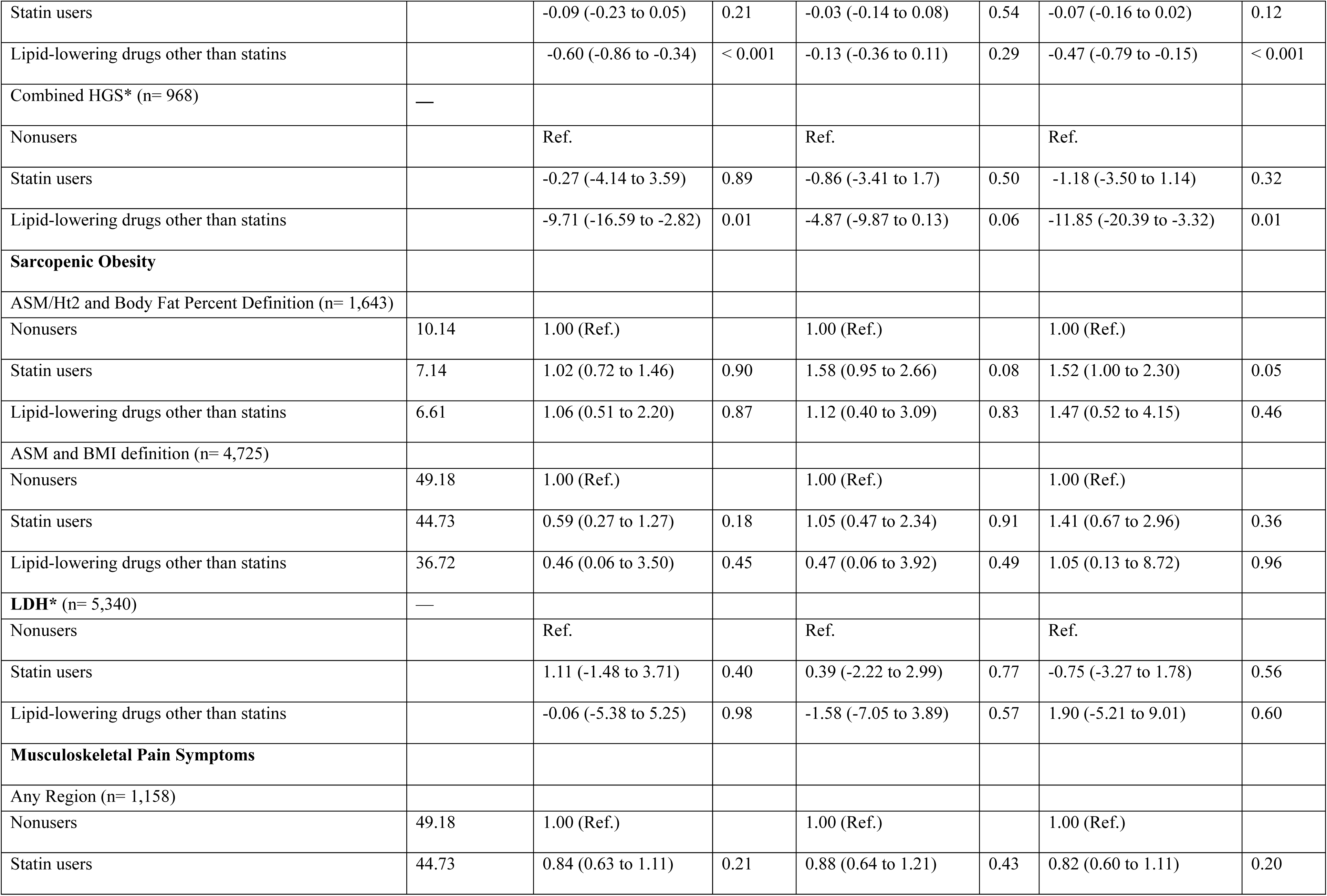
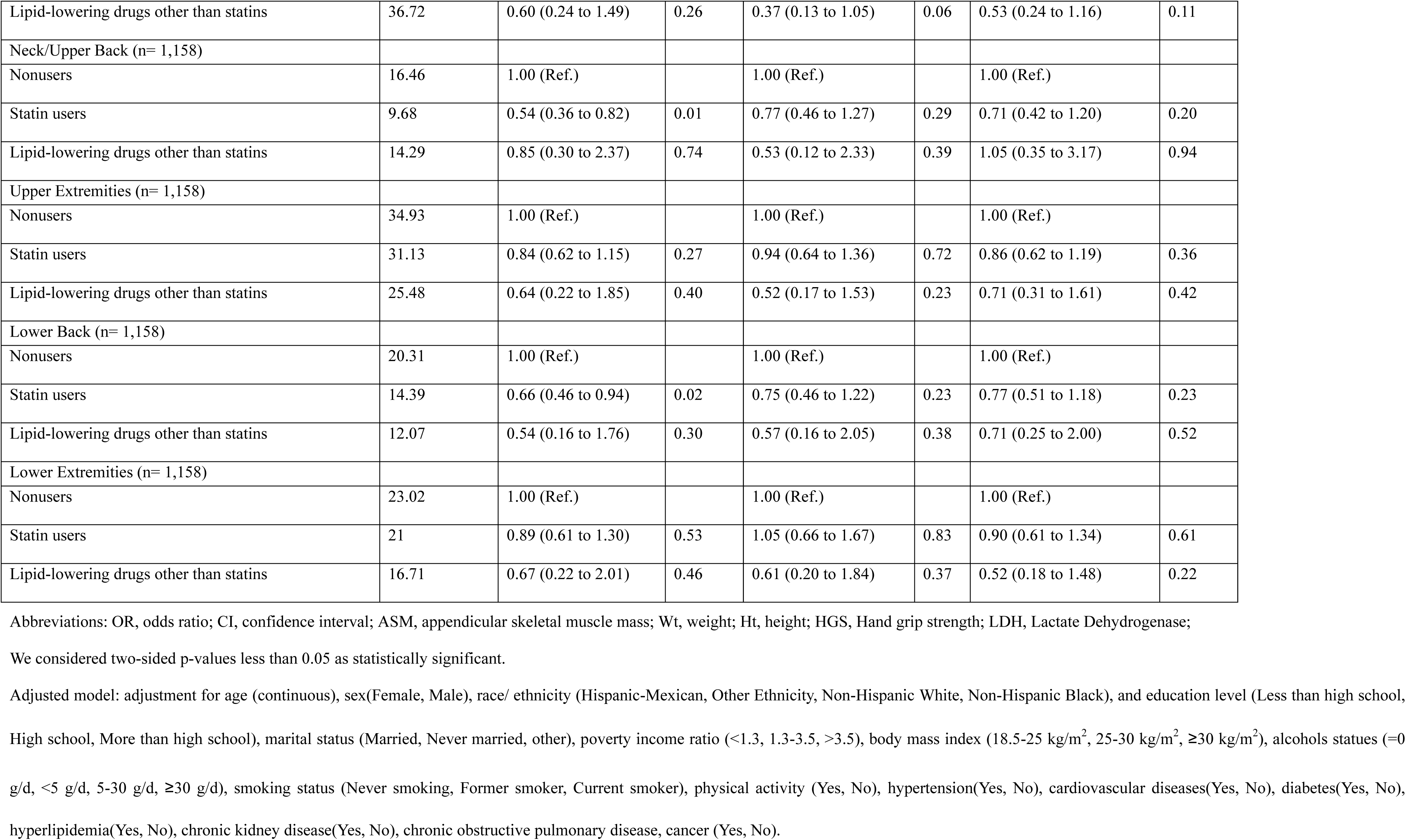
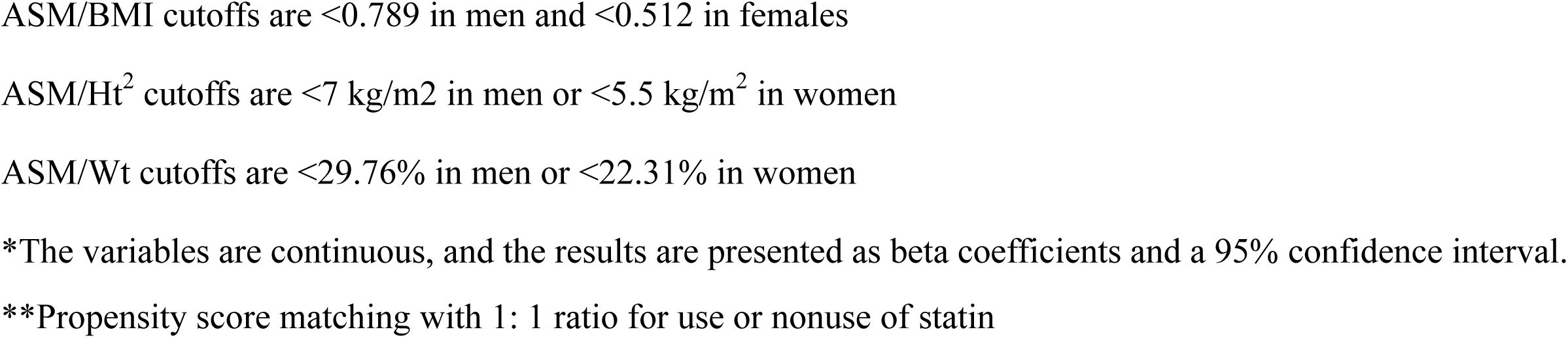
The association between statin users or lipid-lowering users and skeletal muscle-related phenotypes compared with nonusers among participants with cardiovascular disease.

**Table S10.**
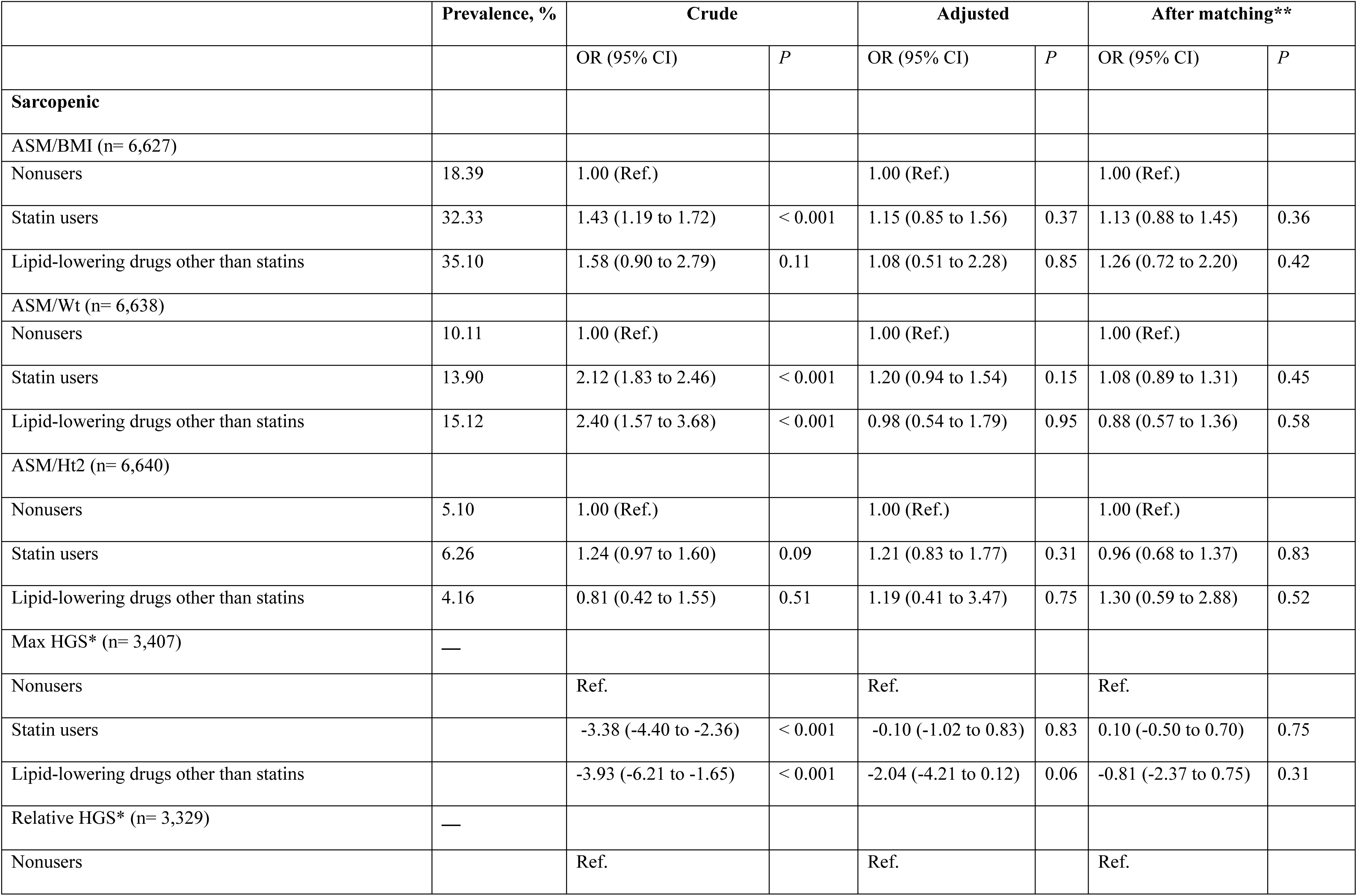
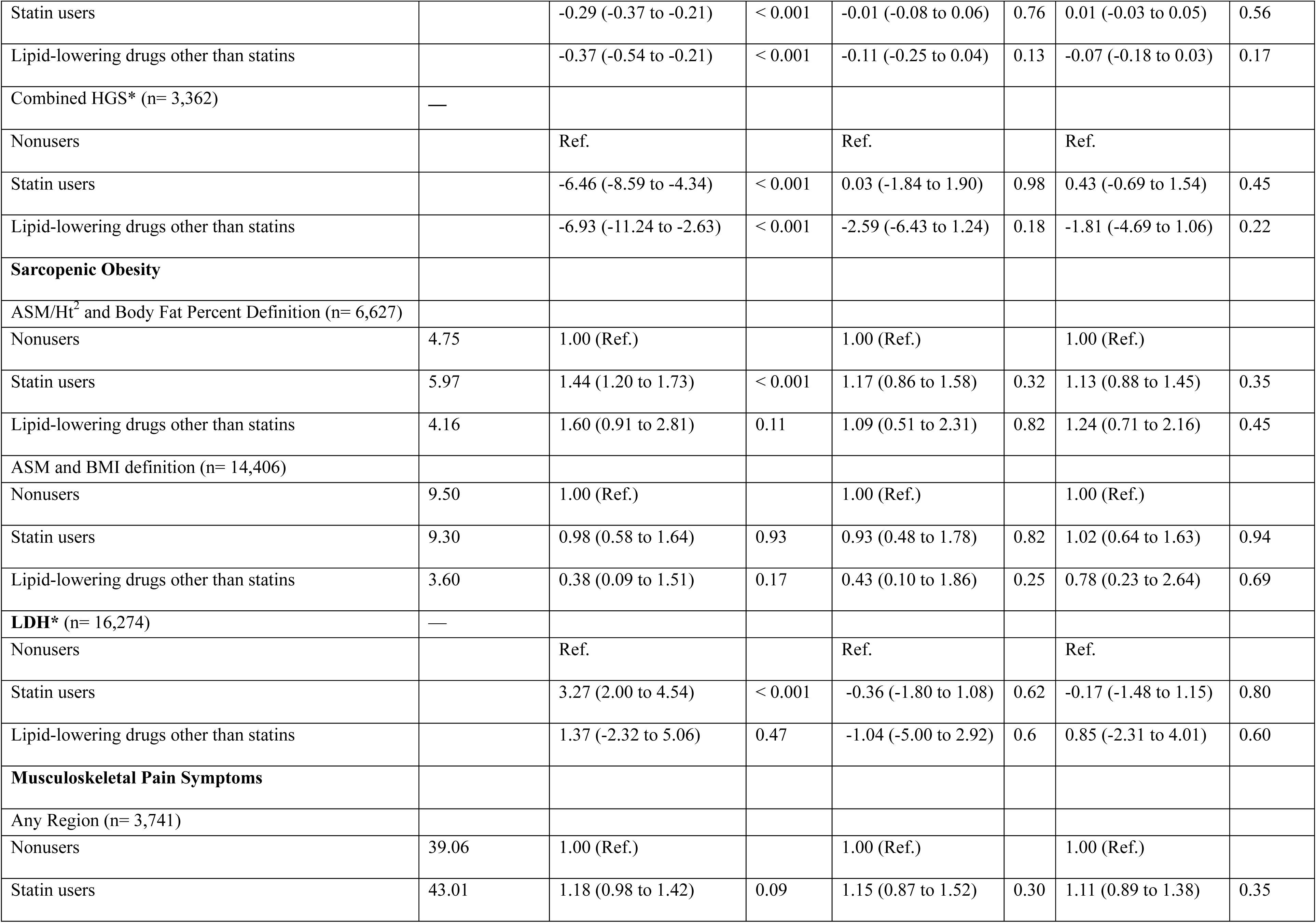
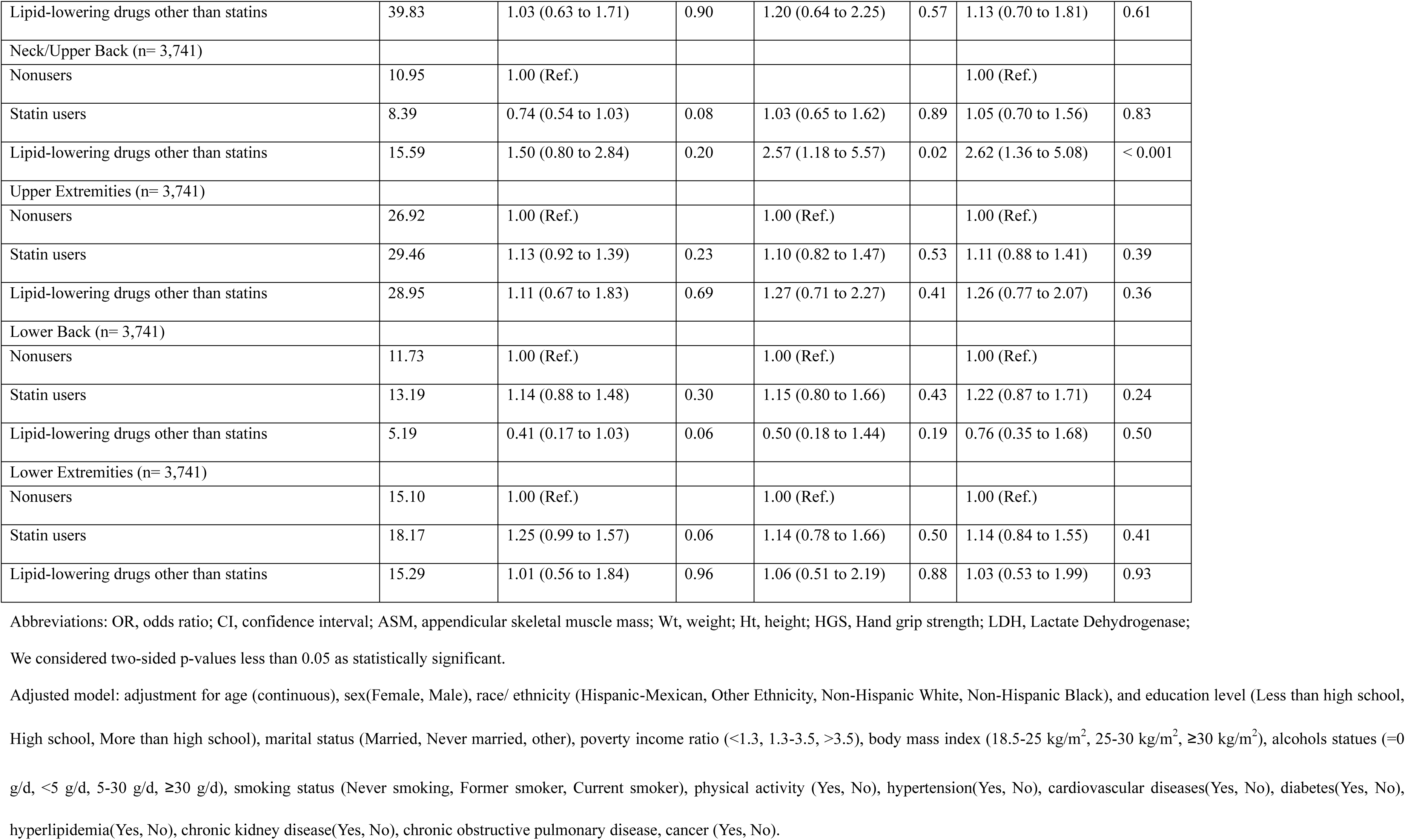
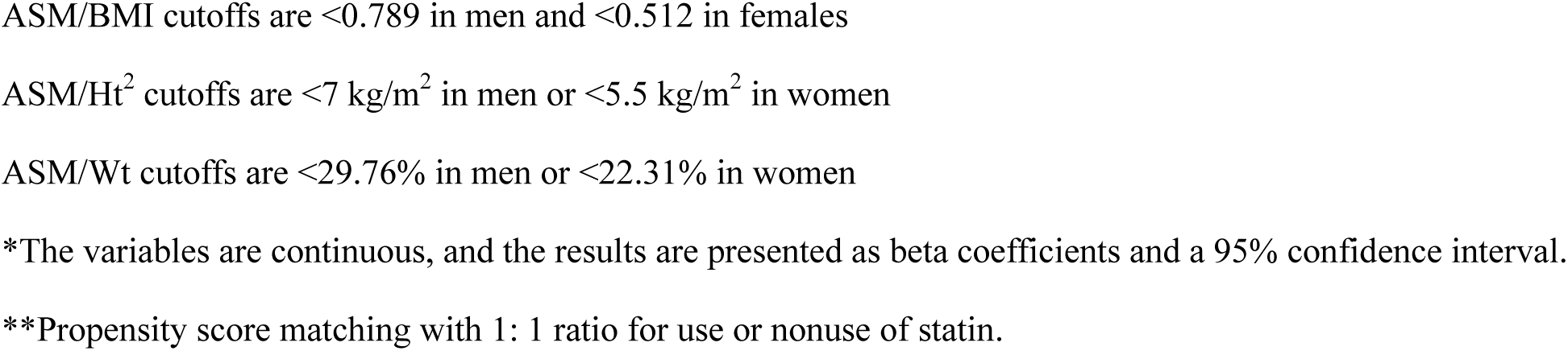
The association between statin users or lipid-lowering users and skeletal muscle-related phenotypes compared with nonusers among participants with hyperlipidemia.

**Table S11.**
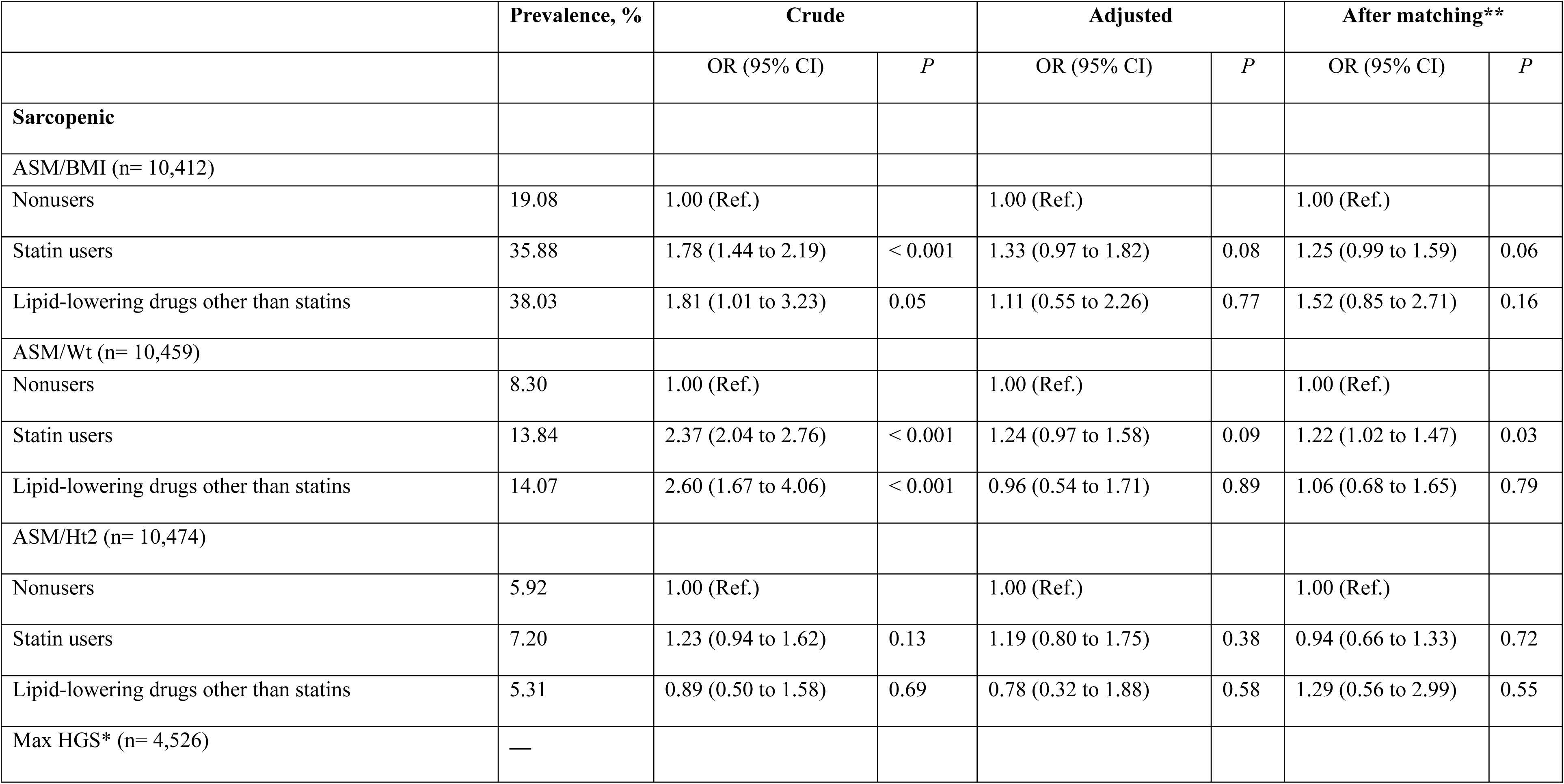
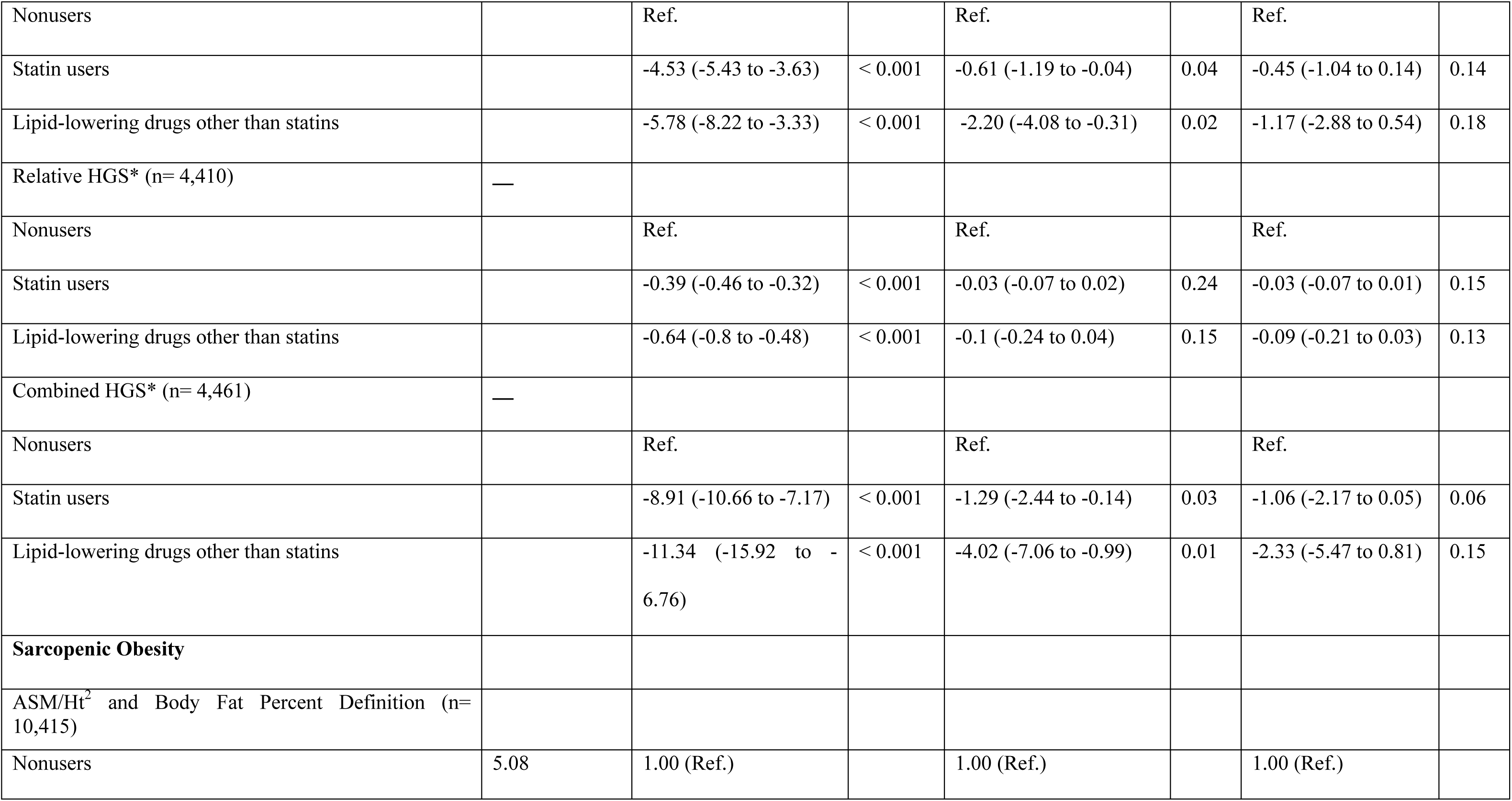
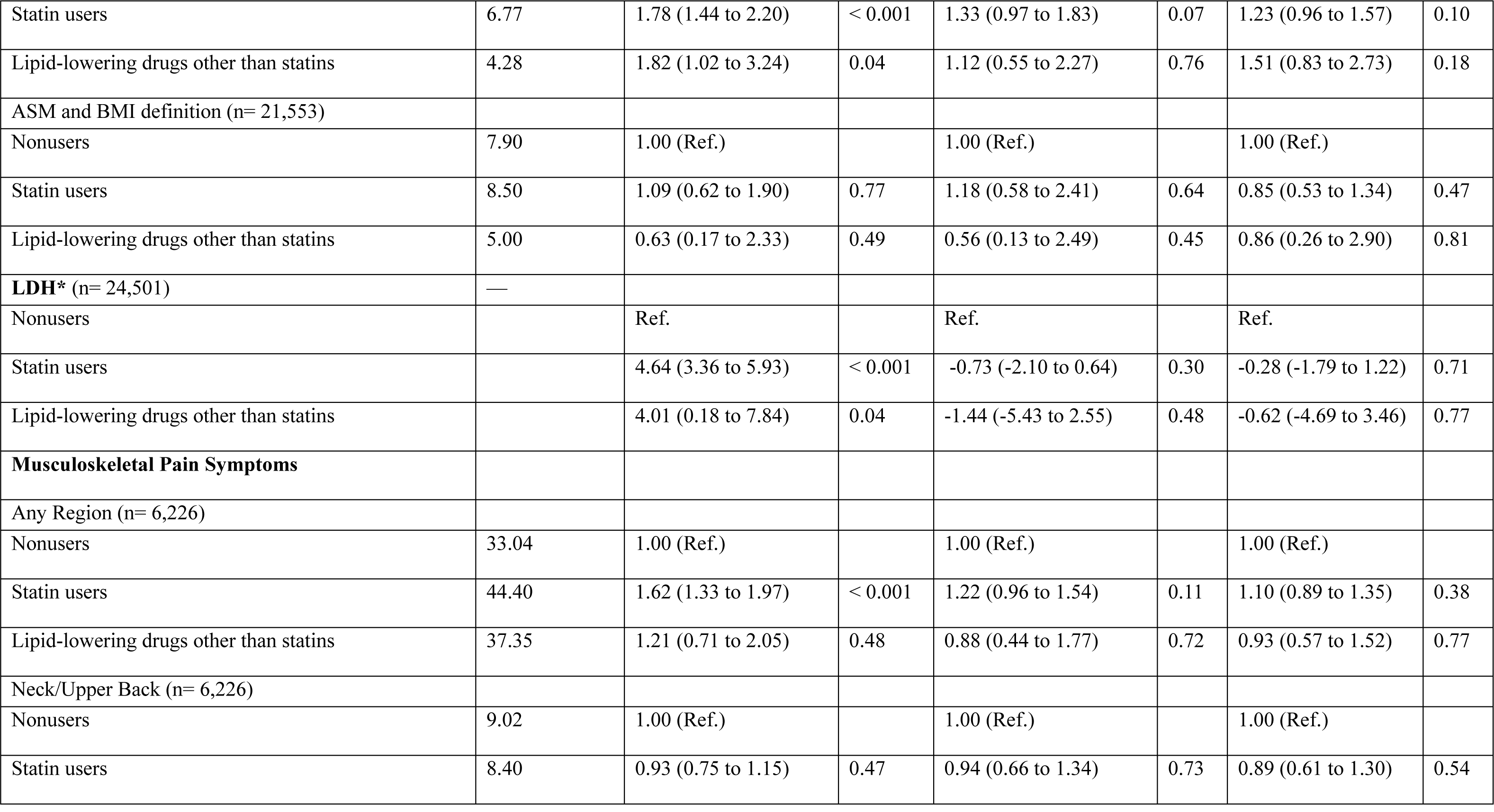
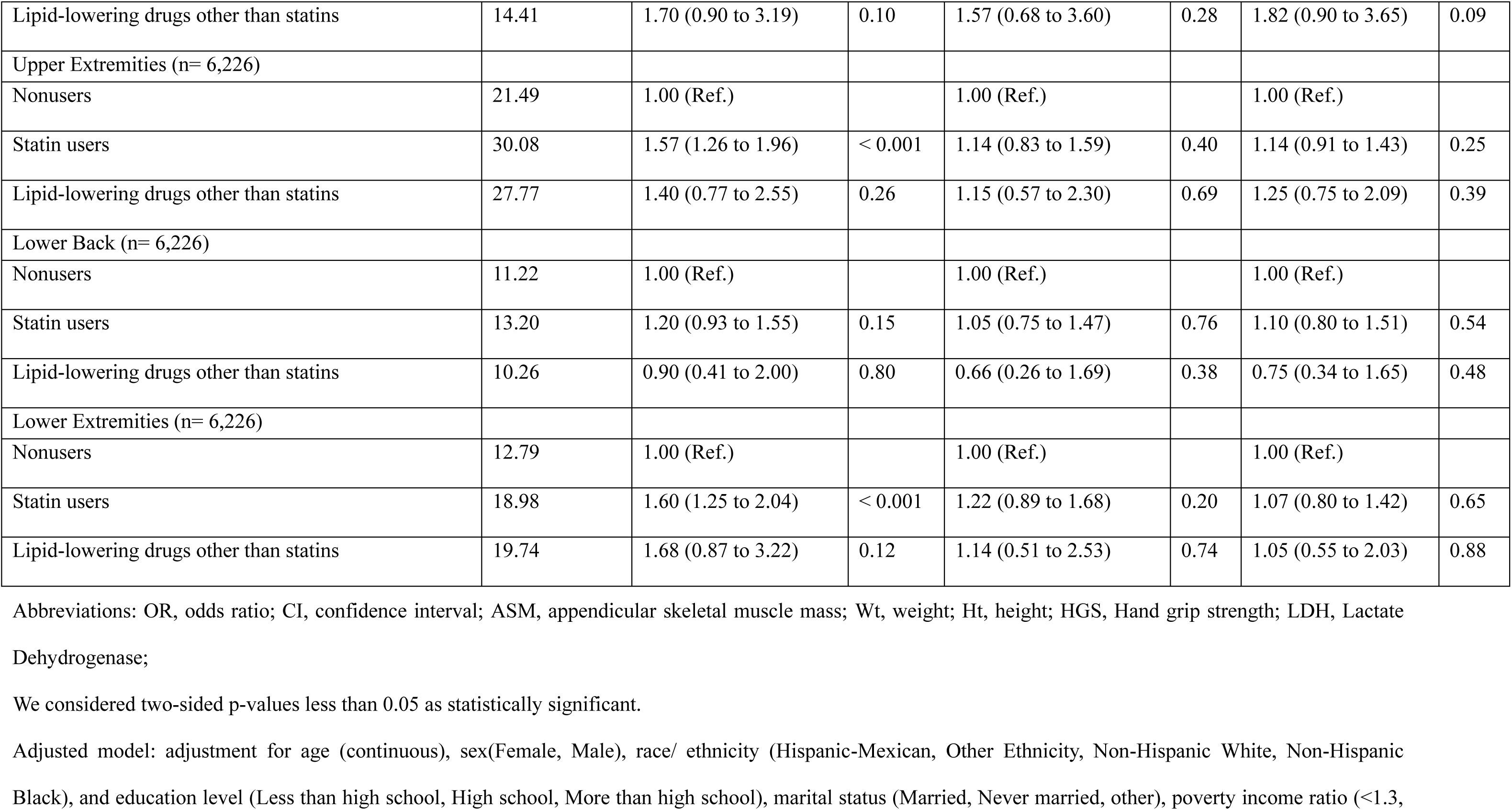
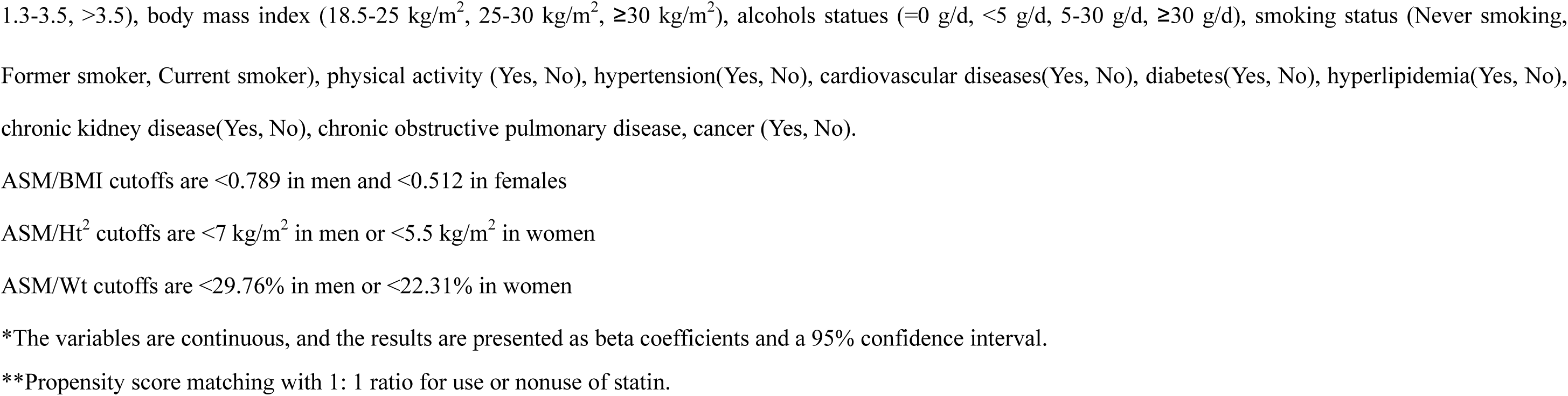
The association between statin users or lipid-lowering users and skeletal muscle-related phenotypes.

